# Reconstruction of a large-scale outbreak of SARS-CoV-2 infection in Iceland informs vaccination strategies

**DOI:** 10.1101/2021.06.11.21258741

**Authors:** Kristjan E Hjorleifsson, Solvi Rognvaldsson, Hakon Jonsson, Arna B Agustsdottir, Margret Andresdottir, Kolbrun Birgisdottir, Ogmundur Eiriksson, Elias S Eythorsson, Run Fridriksdottir, Gudmundur Georgsson, Kjartan R Gudmundsson, Arnaldur Gylfason, Gudbjorg Haraldsdottir, Brynjar O Jensson, Adalbjorg Jonasdottir, Aslaug Jonasdottir, Kamilla S Josefsdottir, Nina Kristinsdottir, Borghildur Kristjansdottir, Thordur Kristjansson, Droplaug N Magnusdottir, Runolfur Palsson, Louise le Roux, Gudrun M Sigurbergsdottir, Asgeir Sigurdsson, Martin I Sigurdsson, Gardar Sveinbjornsson, Emil Aron Thorarensen, Bjarni Thorbjornsson, Marianna Thordardottir, Agnar Helgason, Hilma Holm, Ingileif Jonsdottir, Frosti Jonsson, Olafur T Magnusson, Gisli Masson, Gudmundur L Norddahl, Jona Saemundsdottir, Patrick Sulem, Unnur Thorsteinsdottir, Daniel F. Gudbjartsson, Pall Melsted, Kari Stefansson

## Abstract

The spread of SARS-CoV-2 is dependent on several factors, both biological and behavioral. The effectiveness of various non-pharmaceutical interventions can largely be attributed to changes in human behavior, but quantifying this effect remains challenging. Reconstructing the transmission tree of the third wave of SARS-CoV-2 infections in Iceland using contact tracing and viral sequence data from 2522 cases enables us to compare the infectiousness of distinct groups of persons directly. We find that people diagnosed outside of quarantine are 89% more infectious than those diagnosed while in quarantine, and infectiousness decreases as a function of the time spent in quarantine. Furthermore, we find that people of working age, 16-66 years old, are 47% more infectious than those outside that age range. Lastly, the transmission tree enables us to model the effect that given population prevalence of vaccination would have had on the third wave had they been administered before that time using several different strategies. We find that vaccinating in order of ascending age or uniformly at random would have prevented more infections per vaccination than vaccinating in order of descending age.

## Introduction

Over 160 million cases of SARS-CoV-2 have been diagnosed globally, resulting in over 3.3 million deaths^1^. As the virus spreads, nations have invested heavily in monitoring the epidemic, including tracking the spread by various means, such as the number of persons diagnosed by testing, serological surveys or by tracking deaths due to COVID-19. The first case of SARS-CoV-2 infection in Iceland was confirmed on February 28, 2020 and as of May 14, 2021 a total of 6526 people have been diagnosed in the country. The first wave in Iceland was characterized by several persons introducing the virus from various countries, initially from travelers returning from ski vacations in Italy and Austria^2^. Extensive sequencing of the viral genomes showed that the first wave in Iceland consisted of several genetically distinct outbreaks that were eliminated by May 2020 through non-pharmaceutical interventions, primarily isolation of cases, contact tracing, quarantine, social restrictions, and mandatory testing at the border^2^. The second and third waves arose at the end of July and in mid-September 2020. Although these waves overlapped in time they can be distinguished with the sequences of the viral genomes. The third wave was considerably larger, consisting of 2783 confirmed cases and was characterized by a single genetic clade, traced back to a person who entered the country in August 2020.

Although the outbreaks in Iceland are relatively small by global standards they are well characterized with good availability of data. Every diagnosed case was contact traced and recent contacts placed in quarantine. At the end of quarantine, all persons were tested and allowed to leave quarantine given a negative test result. This exit test allows for diagnosis of asymptomatic cases that otherwise could have gone undiagnosed^3^. In order to aid the contact tracing effort, every sample that tested positive by PCR was sequenced within 36-48 hours of the sample collection and the results fed back to the contact tracing team to inform their inquiries. Furthermore, every infected person was enrolled in telehealth monitoring and received multiple structured phone calls to monitor symptoms after diagnosis and provide support regarding isolation practice^3^.

This extensive data collection can provide insight into the effectiveness of measures taken to curb the spread of the virus. To this end we focused on the third wave of the pandemic in Iceland and gathered all available data to computationally reconstruct the transmission of the virus from person to person. This constitutes the largest study to date investigating a single outbreak with complete contact tracing and sequence data.

Being able to understand the differences between distinct groups of persons in epidemic outbreaks is a key to being able to employ targeted measures to contain them. By reconstructing the chain of events in an entire outbreak, we can observe these differences directly. Furthermore, having access to a case-by-case replay of the outbreak enables us to model the effect that vaccinations would have had on the third wave, had they been administered before that time.

### Outbreak reconstruction

In any outbreak of a viral disease there is a single progenitor, who infects a number of persons, each of whom in turn infects other persons and so forth until the disease is contained or everyone has been infected. These transmissions from person to person form a *tree of transmissions* with the progenitor as its root. Since the third wave in Iceland originated with one infected person entering the country, it consisted mostly of a single subtree of the global transmission tree of the SARS-CoV-2 pandemic. Although other clades were introduced at the border, none of them gained foothold. Despite the extensive data collected on each diagnosed case, the true transmission tree of the third wave cannot be determined from them with certainty. In the contact tracing data, the contact resulting in a transmission may not be reported and there are reported contacts between persons where a transmission did not occur. Even in cases where an actual transmission took place, the direction of transmission is not necessarily known unless the virus accumulated a mutation at transmission (Figure 1.B). Therefore, we extended the *Outbreaker2* model^4,5^ to infer the most likely transmission trees using data from contact tracing, sequences of the viral genome, household membership, and times of onset of symptoms, quarantine, and diagnosis.

**Figure 1.**
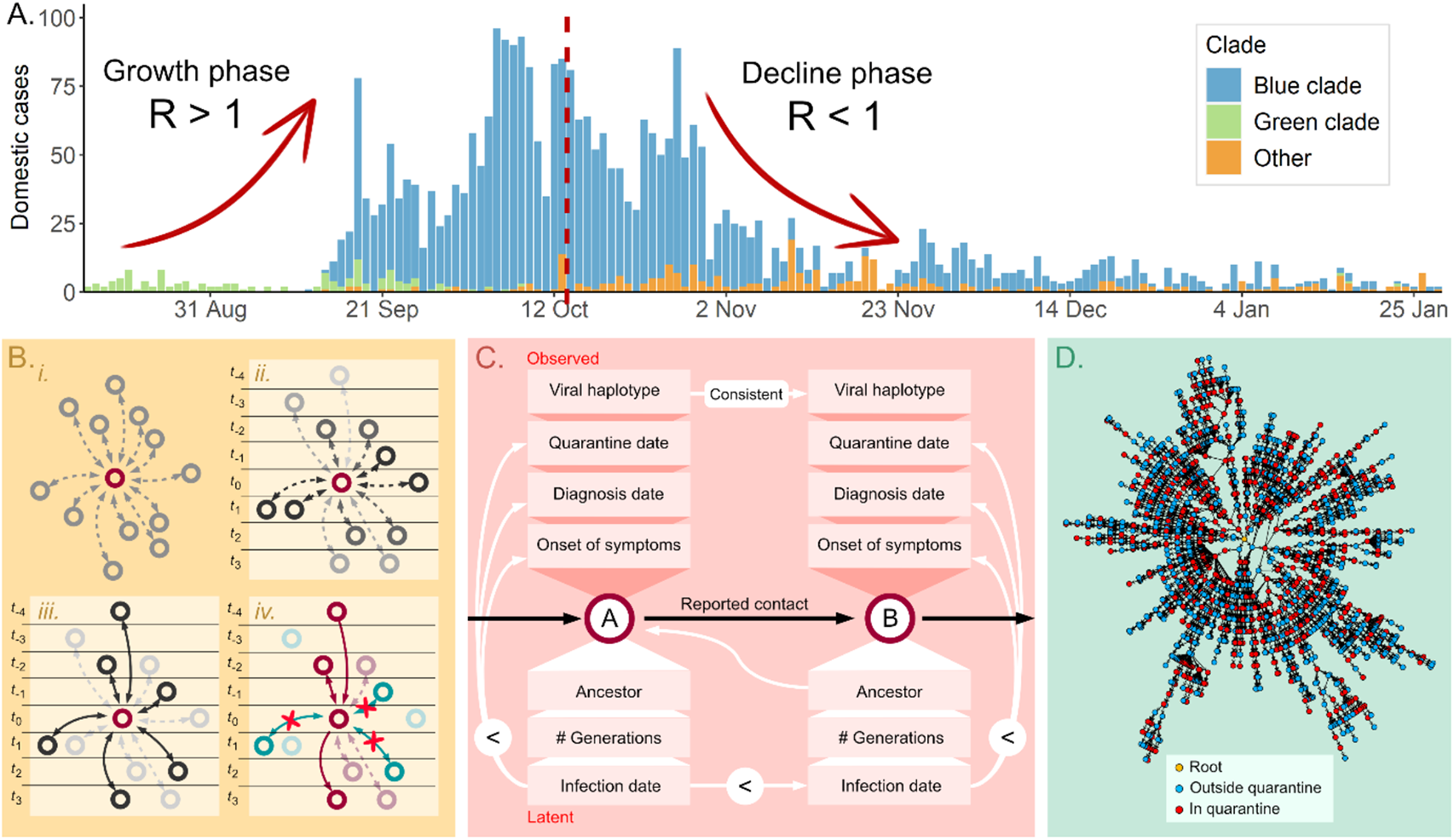
**A**. Daily cases during the third wave of SARS-CoV-2 infections in Iceland, excluding cases diagnosed at the border. The outbreak was in growth until around October 15th, 2020 (red dashed line) and in decline until the end of January 2021. **B**. *i*. When determining who infected a person, initially all diagnosed cases are equally likely. *ii*. Quarantine, diagnosis and dates of symptom onset make some people more likely than others, assuming specific incubation time and generation time distributions. *iii*. Contact tracing data make certain transmissions very likely but do not enable us to disregard others. *iv*. Given the viral haplotypes, we can disregard transmissions where the haplotypes are incompatible, i.e. neither is derived from the other, and in some cases determine the direction of the transmission, in cases where de novo mutations occur between generations. **C**. We use the real-world data and the tree structure to infer the latent data for each diagnosed case. The “<”-symbol represents that the date on the left needs to precede the date on the right. For each diagnosed person we infer the ancestor, i.e. the person who infected them, the date of infection, and the number of transmissions separating the ancestor and the person, *κ*. **D**. One instance of a reconstructed transmission tree for the third wave in Iceland.

### Estimating stratified reproduction number using transmission trees

The *effective reproduction number R* of a disease outbreak denotes how many persons each diagnosed person infects on average. It is a useful metric for discerning whether an epidemic is in growth or whether it is successfully being contained. In order to contain an outbreak, *R* must stay below one. The *R* at a given time is denoted by *R*_*t*_ and is called the *time-varying reproduction number*.

A variety of methods have been proposed to estimate *R*_*t*_, most of them relying on the daily number of cases or deaths and assumptions about incubation time and generation time distributions^6–10^. These methods operate by attributing the number of cases at time *t* to cases diagnosed in the preceding days weighted with the assumed generation time distribution. The idea of reconstructing the latent transmission tree of an outbreak has been explored in previous studies^4,11,12^, most recently with the Outbreaker2 model which infers the transmission tree of an outbreak using contact data, sequence data and times of symptom onset. In comparison to the classical methods, in a transmission tree model, *R*_*t*_ is calculated by averaging the *out-degree*, i.e. the number of persons they infected, of everyone in the tree at time *t*. Furthermore, since the data are available on an individual level, we can estimate the reproduction number for distinct groups of people in the transmission tree, allowing us to compare their relative infectiousness in an outbreak.

In this study, we expand upon the size of transmission trees reconstructed in previous studies by analyzing an entire epidemic on a national scale^4,12^. This gives us the statistical power to quantify the efficacy of targeted interventions such as quarantine measures and compare the infectiousness of different age groups at different times over the course of the epidemic. Current methods do not include in their models much of the information we possess, such as quarantine times, household data and the fact that the third wave in Iceland was a single introduction outbreak. Therefore, we extended the Outbreaker2 model to infer the transmission tree of the third wave (Figure 1.C, Supplementary methods).

### Simulating the effects of vaccination on transmission trees

There are two distinct goals of the vaccination effort. Firstly, to protect those at risk, such as the elderly, those with underlying diseases, and front-line workers. Secondly, to obtain herd immunity to protect the community from future outbreaks. Once the first goal has been attained, the order in which vaccines should be distributed to the rest of the population needs to be decided. Some efforts have been made to simulate the effect vaccination has on the spread of the disease^13–15^. These models construct a theoretical wave of infections assuming a compartmental model (e.g. SIR models and variations thereof), and rely on multiple epidemiological constants. However, by using the transmission trees, we can use real-world data to reconstruct what would have happened if certain people in the tree had been immune at the time of the third wave. By simulating immunity given a specific fraction of the adult population vaccinated, we can estimate what the size of the third wave in Iceland would have been, conditioned on all non-pharmaceutical interventions having been the same. Using this method, we can simulate the effect different vaccine distribution strategies would have had on the third wave, had they been employed before that time.

## Results

In the third wave of SARS-CoV-2 infections in Iceland, 89% of diagnosed cases had a single dominant haplotype, traced back to a person who entered the country in August 2020 (Figure 1.A). The third wave accumulated 2783 diagnosed cases of the same or derived haplotype (colloquially referred to as the *blue clade*, see Supplementary methods) over a period of five months before being contained. The last case was diagnosed at the end of January 2021. Although cases of other clades were diagnosed during this period, we refer to the blue clade outbreak in Iceland as *the third wave* for the sake of brevity. Vaccinations against SARS-CoV-2 were initiated in Iceland on December 28th, 2020 and when the last blue clade case was diagnosed on January 28th, 2021, only 3.6% of the adult population (16 years and older) had received at least one vaccine dose. The success of the containment was largely due to non-pharmaceutical interventions, mass testing and effective contact tracing measures.

We inferred a transmission tree using data on every person in the third wave diagnosed before December 1st, 2020, a total of 2522 people (Figure 1.D). Of these, 2431 had the blue clade and the remaining 91 had an incomplete or missing haplotype but were included because of contact tracing data. Although the third wave continued past December 1st, this dataset contains 91% of the 2783 persons who were ever diagnosed with this clade. Contact tracing data, quarantine status, and onset of symptoms were available for everyone in the dataset. A total of 1275 (51%) people were diagnosed while in quarantine and 1964 (78%) had reported contact with prior cases. The number persons symptomatic upon diagnosis was 1738 (69%) and 187 (7%) never showed any symptoms at all. An average of 303 persons (12%) in the model had a number of transmissions between them and their ancestor greater than one, indicating the presence of undiagnosed cases (Supplementary methods). The Outbreaker2 model estimated the proportion of observed cases to be 87% of the total (95%-PI: 83%-91%) (Supplementary table 5).

### Effect of contact tracing-informed quarantine on the effective reproduction number

Stratifying the diagnosed cases by whether or not they were in quarantine at the time of diagnosis and calculating 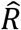, an estimate of the effective reproduction number *R*, for the respective groups gives us a metric for the effectiveness of the contact tracing and government-mandated quarantine employed in Iceland, in the third wave. Those diagnosed outside of quarantine were 88.8% more infectious (95%-CI: 70.9%-109.2%, p=2.8×10^−32^) than those diagnosed while in quarantine. The former had an 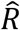 of 1.31 (95%-CI: 1.21-1.43) while the latter had an 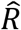 of 0.70 (95%-CI: 0.66-0.73). Furthermore, the length of time from the start of quarantine to a positive PCR test had a significant effect on infectiousness. Persons diagnosed after a short quarantine, i.e. one or two days, were 66.6% more infectious (95%-CI: 49.3%-85.2%, p=4.0×10^−19^) than those diagnosed after a long quarantine, i.e. three or more days, with an 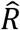 of 0.89 (95%-CI: 0.83-0.96) and 0.54 (95%-CI: 0.50-0.58), respectively. This indicates that the sooner people were quarantined after exposure, the fewer opportunities they had to infect others, which in turn shows that contact tracing is highly time critical. Additionally, those diagnosed outside quarantine were 144.4% more infectious (95%-CI: 116.8%-174.1%, p=2.5×10^−50^) than those who were diagnosed after a long quarantine.

### Effective reproduction number varies with age

By stratifying all diagnosed cases by age, we were able to compare the infectiousness of different age brackets. We calculated the effective reproduction number of adults, 16 years and older, and children, 15 years and younger, demonstrating that adults were 59.5% more infectious (95%-CI: 41.6%-83.4%, p=1.7×10^−12^), with an 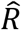 of 1.06 (95%-CI: 0.99-1.12) compared to 0.66 (95%-CI: 0.59-0.73) for children. We also calculated the 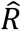 of those of working age, 16 to 66 years old and found that they were 46.6% more infectious (95%-CI: 27.7%-65.4%, p=1.6×10^−8^) than those outside that age range, with an 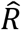 of 1.08 (95%-CI: 1.01-1.16) compared to 0.74 (95%-CI: 0.66-0.84) for children and the elderly. In addition to showing that adults were more infectious than children and the elderly in the third wave in Iceland, this indicates that people of working age in particular played a key role in the transmission of the virus.

### Estimating the time-varying reproduction number

The effective reproduction number is not constant over the course of an outbreak, from its propagation to its eventual extinction. We calculated 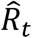, an estimate of the time-varying reproduction number *R*_*t*_, stratified by whether or not people were in quarantine at the time of diagnosis (Figure 2.A). 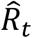 outside quarantine is more variable, than the relatively stable 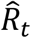 in quarantine. Figure 2.A shows three peaks in 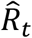 outside of quarantine, which correspond to three well characterized events: two superspreading events and one outbreak in a hospital. On October 15, 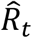 went below 1 outside of quarantine for the first time and stayed below 1 except for the time period covering the hospital outbreak. Based on this observation we split the outbreak into a growth and decline phase on October 15 (Figure 1.A).

**Figure 2.**
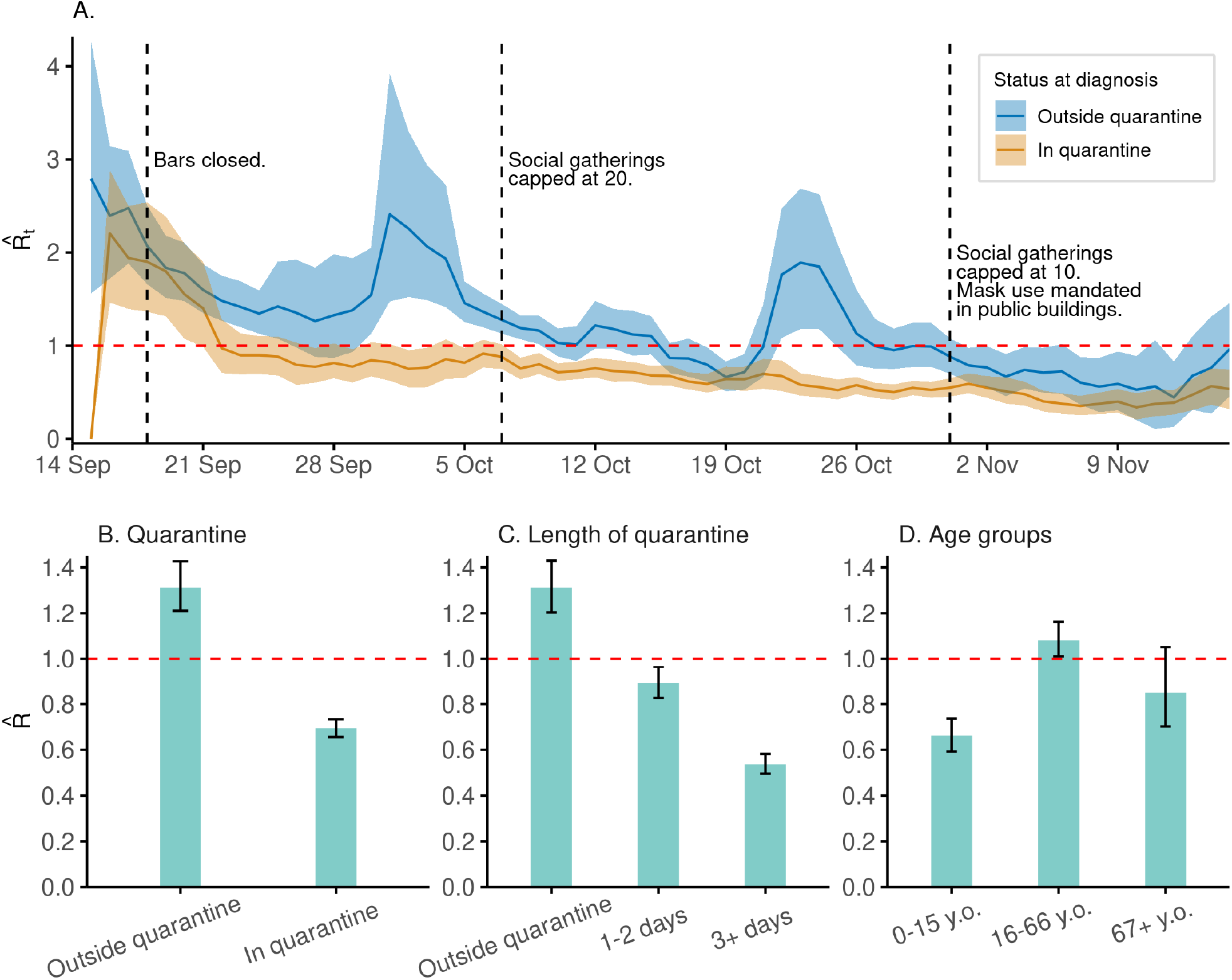
**A**. 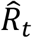for those diagnosed while in quarantine and those diagnosed outside quarantine, respectively. The shaded area represents the 95% confidence interval for the mean and dashed lines show dates of social restrictions imposed. **B**. Effective reproduction number of those diagnosed outside quarantine compared to those diagnosed in quarantine. Error bars reflect 95% CI of the mean. **C**. Effective reproduction number of those diagnosed outside quarantine, those diagnosed after 1-2 days in quarantine and those diagnosed after 3+ days in quarantine. **D**. Effective reproduction number stratified by age.

### An outbreak has (at least) two phases

Any outbreak has at least one growth phase and at least one decline phase. The mean out-degree of people who get infected during a growth phase is strictly greater than one, and the mean out-degree for people who get infected during a decline phase is strictly less than one. Note that for an entire outbreak, the mean out-degree is equal To 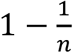, where *n* is the number of people in the transmission tree (Supplementary methods).

The estimated effective reproduction number for different groups during the decline and growth phase of the third wave are shown in Table 1. The growth phase of the third wave in Iceland had an 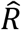 of 1.17 (95%-CI: 1.09-1.27) and the decline phase had an 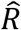 of 0.77 (95%-CI: 0.70-0.86). All comparisons reported above remain significant in the growth phase and the decline phase, except there is not a significant difference between the infectiousness of those of working age and the infectiousness of those outside working age in the decline phase (23.8%, 95%-CI: -3.3%-56.3%, p=0.08). Full results of the comparisons can be found in Supplementary table 1.

**Table 1.** The number of people in different groups diagnosed in the growth phase and the decline phase of the third wave of SARS-CoV-2 infections in Iceland, and their estimated effective reproduction number 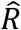.

| Group | Overall |  | Growth phase |  | Decline phase |  |
| --- | --- | --- | --- | --- | --- | --- |
| | N | $\hat{R}$ | N | $\hat{R}$ | N | $\hat{R}$ |
| Everyone | 2522 (100%) | 1.00 | 1442 (100%) | 1.17 (1.09-1.27) | 1080 (100%) | 0.77 (0.70-0.85) |
| Outside quarantine | 1247 (49%) | 1.31 (1.21-1.43) | 776 (54%) | 1.45 (1.32-1.62) | 471 (44%) | 1.08 (0.93-1.25) |
| In quarantine | 1275 (51%) | 0.69 (0.66-0.73) | 666 (46%) | 0.84 (0.78-0.91) | 609 (56%) | 0.53 (0.49-0.57) |
| Short quarantine | 564 (22%) | 0.89 (0.83-0.96) | 340 (24%) | 1.02 (0.93-1.13) | 224 (21%) | 0.70 (0.62-0.78) |
| Long quarantine | 711 (28%) | 0.54 (0.50-0.58) | 326 (23%) | 0.66 (0.58-0.74) | 385 (36%) | 0.43 (0.39-0.48) |
| Adults (16+ years old) | 2164 (86%) | 1.06 (0.98-1.12) | 1269 (88%) | 1.22 (1.13-1.32) | 895 (83%) | 0.82 (0.74-0.91) |
| Children (0-15 years old) | 358 (14%) | 0.66 (0.59-0.73) | 173 (12%) | 0.80 (0.69-0.93) | 185 (17%) | 0.53 (0.45-0.62) |
| Working age (16-66 years old) | 1921 (76%) | 1.08 (1.01-1.16) | 1171 (81%) | 1.25 (1.15-1.37) | 750 (69%) | 0.82 (0.73-0.92) |
| Outside working age | 601 (24%) | 0.74 (0.66-0.84) | 271 (19%) | 0.83 (0.74-0.92) | 330 (31%) | 0.66 (0.54-0.81) |

### Simulating vaccination strategies

The effect of vaccination is not only determined by the proportion of persons vaccinated, but also who is vaccinated. As our results show, there was a significant difference in infectiousness between age groups in the third wave. To investigate this effect, we modeled three vaccination strategies on the adult population, 16 years and older: vaccinating by order of descending age, order of ascending age, and uniformly at random. We then estimated what the size of the third wave would have been for different levels of vaccination. For each strategy we iteratively increased the proportion of the adult population vaccinated, both starting at 0% and assuming a starting point of 29% as of April 28th, 2021. The second starting point reflects the actual vaccinations of persons at high risk and frontline workers in Iceland. We assumed that a single dose would lower the probability of being infected by 60% and that two doses would lower it by 90%^16–18^.

Figure 3 shows the mean size of the outbreak for the three vaccination strategies, assuming the first person in the transmission tree is unvaccinated. As a benchmark we compare the vaccination strategies by the lowest proportion of adults who would have needed to be vaccinated such that the final size of the third wave would have been 100 persons (4% of the observed outbreak) on average. These simulations are not sensitive to the initial cases in the transmission tree (Supplementary methods, supplementary figures 1-4).

**Figure 3.**
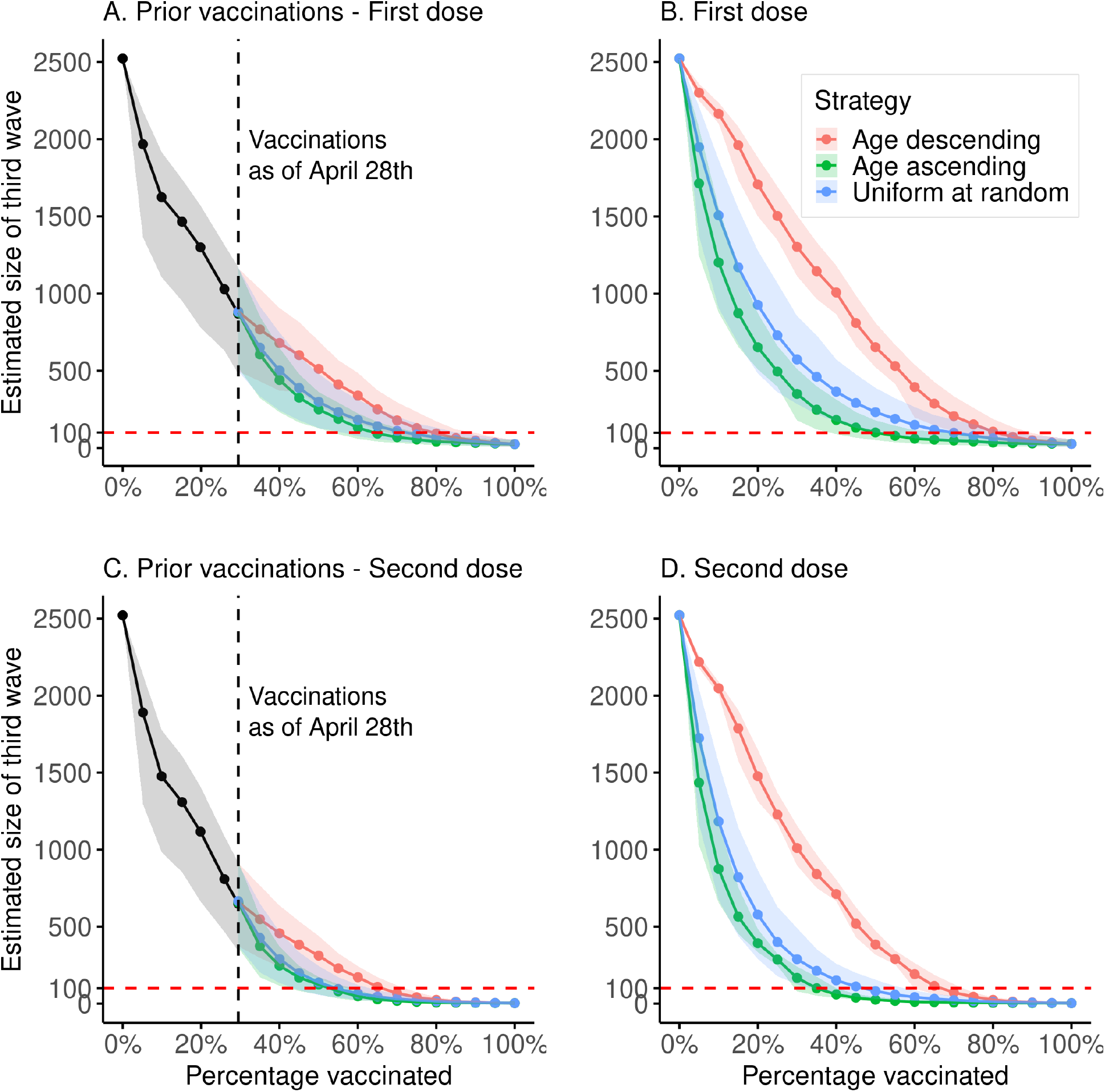
Simulations of the estimated final size of the third wave at a given population prevalence of vaccination. Solid lines show the mean size of the outbreak, shaded areas represent 2.5%-97.5% quantiles. **A**. Using the actual vaccination scheme for at-risk groups and front-line workers, up to 29% of the adult population, and using three separate vaccination strategies from 29% to 100%: age-descending, age-ascending and uniformly at random. Modeled vaccinations beyond the 29% mark are assumed to have an efficacy of 60%. **B**. Simulations of the size of the third wave, assuming 60% vaccine efficacy, under the three different vaccination strategies, starting with no vaccinations and concluding with 100% of the adult population vaccinated. **C**. Same simulation as in A, but all vaccinations are assumed to have an efficacy of 90% (both first and second dose administered). **D**. Same simulation as in C, but assuming 90% vaccine efficacy.

All non-pharmaceutical interventions being the same as they were in the third wave, starting at 29% and vaccinating with a single dose in order of descending age would have yielded an outbreak with a mean size of 100 persons with 79% of adults vaccinated (95%-CI: 68%-89%). Vaccinating in order of ascending age would have yielded a 100 person outbreak with 64% of adults vaccinated (95%-CI: 54%-76%), and vaccinating uniformly at random with 72% vaccinated (95%-CI: 56%-85%). Table 2 shows comparisons between the different vaccination strategies.

**Table 2.** The lowest proportion of adults who would have needed to be vaccinated such that the final size of the third wave would have been 100 persons on average. The former two models use actual vaccination numbers up to the 29% mark and extrapolate from there using the three strategies. The latter two models start from zero.

| Model | Proportion of adults vaccinated |  |  |
| --- | --- | --- | --- |
|  | Age, descending | Age, ascending | Uniform at random |
| Actual vaccinations/First dose | 79.2% (67.6%-89.4%) | 64.1% (53.7%-75.7%) | 72.3% (56.1%-85.1%) |
| Actual vaccinations/Second dose | 66.2% (57.1%-72.1%) | 52.8% (42.4%-58.4%) | 54.5% (43.7%-63.1%) |
| First dose | 81.1% (71.8%-89.6%) | 50.4% (38.5%-69.2%) | 70.0% (50.0%-86.5%) |
| Second dose | 66.8% (59.9%-72.4%) | 35.0% (29.4%-40.1%) | 47.0% (33.6%-57.5%) |

## Discussion

The spread of the SARS-CoV-2 is dependent on several factors, both biological and behavioral. The effectiveness of various non-pharmaceutical interventions employed by nations can largely be attributed to changes in human behavior. Quantifying this effect remains challenging. By leveraging the extensive data collected for diagnosed persons in the third wave of SARS-CoV-2 infections in Iceland, we created a model that allowed us to observe the differences in infectiousness of distinct groups of people.

Quarantine has been assumed to slow the spread of infectious diseases but the extent to which it is effective has been difficult to quantify because it requires data on the individual level. We found that mandated quarantine significantly decreased the spread of the third wave of SARS-CoV-2 infections in Iceland, with persons diagnosed outside of quarantine being 89% more infectious than those diagnosed while in quarantine. Furthermore, we observed that contact tracing is time critical, by comparing the infectiousness of people diagnosed after one or two days in quarantine to that of those diagnosed after three or more days in quarantine. Lastly, we found that people of working age played a key role in the generation of the third wave in Iceland.

Even though the data collected are extensive there are always infections that will not be diagnosed. Serological measurements following the first wave of SARS-CoV-2 infections in Iceland^3^ estimated that diagnosed cases were 56% of the total and another 14% were quarantined but undiagnosed. Due to the quarantine exit test and higher availability of PCR tests, we expect that at least 70% of the cases in the third wave were diagnosed and therefore included in the transmission tree. Outbreaker2 estimated that 87% of cases that were diagnosed, but this estimate does not include undiagnosed persons who did not infect others.

The effect of vaccination on the spread of the disease has been studied with classical modeling approaches, based on SIR models and variations. These models can yield insights, but a significant uncertainty remains as to how the contacts should be modeled, dependency between age of contacts and variability due to superspreading events. By reconstructing the third wave from real-world data we have circumvented these limitations by removing the behavioral modeling assumptions and simulating the vaccinations directly on the transmission tree.

We found that vaccinating persons in ascending order of age or uniformly at random would have prevented more transmissions per vaccination than vaccinating in descending order of age, in the third wave of SARS-CoV-2 infections in Iceland. Vaccine efficacy is different for different vaccines. Our estimates of the final size of the third wave are sensitive to the assumed vaccine efficacy. However, the relative difference between the modeled vaccination strategies is independent of the efficacy. Furthermore, recent studies suggest that vaccinated persons who become infected have a lower viral load^19^ and may therefore be less likely to infect others. This is not taken into account in the model presented here.

It is important to note that vaccination of a population serves two distinct purposes: firstly, to prevent death and severe illness in groups at high risk, and secondly to curb the spread of the virus in the population. Our results could inform the strategy for vaccinating the remaining adult population once the high-risk groups have been vaccinated. However, while our results for the third wave indicate that vaccinating in ascending order of age would have curtailed the outbreak sooner, this reflects the age composition of this particular outbreak. Vaccinating the remaining adult population uniformly at random is a more robust strategy, since it removes the dependency between who is vaccinated and their age.

The third wave in Iceland was contained using non-pharmaceutical interventions, primarily contact tracing, quarantine, and social restrictions. The vaccination simulations show what would have happened if a given proportion of the adult population had received vaccinations, given the same social restrictions. When interpreting the results, it is important to keep in mind that they only provide a lower bound on the so-called herd immunity threshold.

## Data Availability

All sequences used in this analysis are available in the European Nucleotide Archive (ENA) under accession number PRJEB44803 (https://www.ebi.ac.uk/ena/browser/view/PRJEB44803)
Source code for model construction and analysis is available at
https://github.com/DecodeGenetics/COVID19_reconstruction_iceland .

## Methods

### Contact tracing of SARS-CoV-2 infections

Everyone who tested positive for SARS-CoV-2 was contacted by a team designated by the authorities to track their infection. Positive persons were required to isolate and everyone with whom they had been in contact, within 48 hours of the onset of symptoms was required to go into quarantine. If the place of quarantine was shared with an infected person, the length of quarantine was two weeks, but otherwise a weeklong quarantine was sufficient. At the end of quarantine, they were tested for SARS-CoV-2 regardless of whether they had developed symptoms or not.

### Sequencing

The viral genomes of all positive PCR samples were sequenced at deCODE genetics. We performed reverse transcription and multiplex amplicon PCR on the basis of information provided by the Artic Network initiative (https://artic.network/) to generate complementary DNA and sequencing libraries. Samples were sequenced using either Illumina (n=1939) or ONT (n=1184) technologies. These numbers include cases diagnosed at the border. Illumina sequencing was performed using MiSeq sequencers (MiSeq v2 reagent kits) with 2 x 150 cycle paired-end reads, with up to 48 multiplexed samples per run. Samples for ONT were multiplexed using native barcodes and sequenced using either GridION or PromethION flowcells, version R.9.4.1. Details regarding sequencing are provided in the Supplementary methods.

### Analysis of sequence data

We aligned amplicon sequences to the reference genome of the SARS-CoV-2 (GenBank number, NC_045512.2)^20^. For Illumina sequences we used the latest Burrows– Wheeler Aligner (BWA-MEM) and variants were called with sequencing utilities bcftools^21^ as previously described^2^. For ONT sequences sequence alignment and variant calling was performed using the Artic Network pipeline (https://artic.network/) with default parameters.

### Extending the Outbreaker model

We used the Bayesian phylogenetic model Outbreaker2^4^ to infer the transmission tree in the third wave. This model infers the posterior distribution of the transmission tree with iterative MCMC sampling, generating 10,000 trees in each chain. Four MCMC chains were run and every 50 samples were extracted with a burn-in of 5,000. The likelihood in the model is the product of the genetic, contact, generation time, incubation time and reporting likelihoods. In order to make use of our extensive data on each diagnosed case we implemented custom likelihood functions for the generation time likelihood and the incubation time likelihood, assuming the same distributions as in Flaxman et al., but truncated them with respect to quarantine, onset of symptoms, and diagnosis dates (Supplementary methods). Furthermore, we implemented a custom genetic likelihood function in terms of variations from the blue clade. We estimated the mutation rate of SARS-CoV-2 in the third wave in Iceland (Supplementary methods) and used it as a fixed parameter in the model. This enabled us to control the likelihood of back mutations in the model and to impute missing or incomplete viral haplotypes (Supplementary methods).

### Simulating vaccination strategies

With our collection of likely infection trees, we simulated different vaccination strategies by selecting people 16 years and older to be immune in each respective infection tree and removing them and all their downstream transmissions from that tree. By counting the number of people left in the tree, we obtained a measure of the size of the third wave in Iceland given that a particular fraction of the population would have been vaccinated at the time, and all non-pharmaceutical interventions being identical.

This replay of the outbreak assumes that all transmissions remain the same, except some persons have been immunized and therefore break the chain of transmission, reducing the size of the outbreak.

We considered three distinct vaccination strategies: vaccinating in order of descending age, in order of ascending age, and uniformly at random. We simulated these strategies by first choosing the fraction of the adult population to be vaccinated. Depending on the strategy, we calculated the probability of each person in the transmission tree being vaccinated, based on age and in turn we simulated immunity to infection for vaccinated persons based on the efficacy of the vaccine. Given the simulated set of immune persons, we computed the final outbreak size for each likely transmission tree, where immune persons break the chain of transmission.

The adult population was segmented into ten-year age brackets with each person in a given bracket assumed to be vaccinated with equal probability. We performed 1,000 simulations, sampling immune persons in each iteration and then calculating the average outbreak size over all transmission trees. The outbreak size point estimates were obtained by averaging the mean outbreak size over all simulations and 95% confidence intervals by taking the 2.5% and 97.5% quantiles. A person in a given simulation was selected to be vaccinated based on how large a proportion of their respective age group was vaccinated in the simulation, and a given vaccine efficacy, which was assumed to be 60% for the former dose and 90% for the latter, which is in line with reported efficacy of the mRNA vaccines^16–18^. The number of people in each respective age group was obtained from census data from the Icelandic Registry Office and is accurate as of January 1st, 2021.

We simulated the effect the actual distribution of vaccines in at-risk groups and front-line workers would have had on the third wave. We obtained the de facto vaccination numbers for each age bracket from the Icelandic Directorate of Health (Supplementary tables 3-4). These consisted of the number of people vaccinated per day from the first day of vaccination on December 28, 2020, until at-risk groups and front-line workers had been fully vaccinated, at which point 29% of the adult population had been vaccinated. For each day we modeled the third wave with the actual number of accumulated vaccinations per age bracket.

In order to assess the sensitivity of the simulations to the initial cases in the tree, we repeated the simulations conditioned on the first 50 persons being unvaccinated, and furthermore, we ran the simulations on subtrees of size greater than 100 whose direct ancestor was one of the first 50 persons to be infected (Supplementary methods, supplementary figures 1-4).

### Statistical analysis

In order to estimate the effective reproduction number *R* of a particular group of diagnosed cases, we averaged the out-degree of people in the group in each transmission tree and took the average over all trees. The confidence intervals were calculated by iteratively calculating 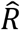 with bootstrapping of the persons in the data set. To estimate the effect size of the difference in infectiousness between two distinct groups of diagnosed cases, we calculated the ratio of 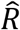 between the groups. Significance was tested by taking the difference of the logs of the bootstrapped 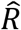 values for the two groups and performing a z-test, using the bootstrapped values to estimate the standard deviation. In addition to bootstrapping, we performed jackknife and permutation tests, with identical results.

We estimated the stratified time-varying reproduction number *R*_*t*_ to be the mean out-degree per group of everyone diagnosed in a four-day sliding window (*t* − 4, *t*], such that each person contributed to 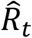 on four distinct days. We obtained the 95% confidence interval with bootstrapping.

## Data and source code availability

All sequences used in this analysis are available in the European Nucleotide Archive (ENA) under accession number PRJEB44803 (https://www.ebi.ac.uk/ena/browser/view/PRJEB44803)

Source code for model construction and analysis is available at https://github.com/DecodeGenetics/COVID19_reconstruction_iceland

## Study oversight

The study was approved by the National Bioethics Committee of Iceland (approval no. VSN-20-070), after review by the Icelandic Data Protection Authority (DPA).

