## Supplementary Appendix for "Reconstruction of a large-scale outbreak of SARS-CoV-2 infection in Iceland informs vaccination strategies"

### Supplementary Methods

#### Extended Outbreaker2 model for COVID-19 outbreak in Iceland

To reconstruct the tree of transmissions (who infected whom) in the third wave of COVID-19 in Iceland, we extended the Outbreaker model as implemented in the Outbreaker2 R package. Outbreaker2 is a Bayesian phylogenetic model framework, which infers transmission trees given temporal, genetic and contact data of infected persons. By Bayes' theorem, the posterior probability of a transmission tree is proportional to the product of its prior probability and the likelihood of the data under it. The likelihood itself is the product of five components, incubation time, infection time delay, contact information, viral haplotype and reporting of infections. The R package is flexible in that the user can implement their own likelihood functions for each of the five components. Making use of the extensive data available, we extended the sampling time, infection time delay and genetic likelihoods as described below. The output of the model is a sequence of transmission trees, which can be thought of as independent samples of the posterior distribution of transmission trees conditioned on the data.

##### Incubation time likelihood

The incubation period of an infectious disease is the time from exposure to onset of symptoms. Because of the extensive follow-up of each person diagnosed with COVID-19 (see Completeness of symptom and contact data, Supplementary methods), the date of symptom onset in our data is well recorded. Furthermore, in instances where persons were in quarantine at the time of diagnosis, we can constrain a sample from the incubation time distribution to be upper bounded by the quarantine date, conditioned on that they are not in the same household as an infected person. For symptomatic persons, the incubation distribution, $f^*$, was chosen to be gamma distributed, like in Flaxman et al[7]

$$f^* \sim \Gamma(\alpha = 1.35, \beta = 0.27)$$

discretized by taking the density at each $t \in [1, \cdots, 100]$ and dividing by the sum over all $t$. For asymptomatic persons, a uniform distribution was chosen

$$f(t) = \frac{1}{14}, t = 1, \ldots, 14$$

For each person, the incubation distribution was constrained based on the available data in the following way. Let $t_{r,i} = \min\{t_{d,i}, t_{s,i}\}$ where $t_{d,i}$ is the time of diagnosis, $t_{s,i}$ is the time of symptom onset (if not reported, $t_{s,i} = \infty$). We define an upper bound on the infection time of person $i$ to be $t_{u,i} = min\{t_{r,i}, t_{q,i} + 1\}$ where $t_{q,i}$ is the quarantine time (if $i$ is not quarantined or is in a household with infected persons, then $t_{q,i} = \infty$ ). The likelihood $\mathcal{L}_{TI}$, of the infection time of person $i$, $t_{inf,i}$, is:

$$\mathcal{L}_{TI}(t_{inf,i}) = \begin{cases} 0, & \text{if } t_{inf,i} > t_{u,i} \\ \dfrac{f(t_{r,i} - t_{inf,i})}{\sum_{k > t_{r,i} - t_{u,i}} f(k)}, & \text{otherwise.} \end{cases}$$

Finally, we lower bound the infection time of all persons at seven days before the first date of diagnosis in the third wave.

#### Generation time likelihood

Generation time is the time from one transmission to the next in a chain of infections. Since the infection times are mostly unknown, this was approximated with the serial interval distribution, i.e. the time between onsets of symptoms of one to the next in a chain of infections. As with the incubation time distribution, we can place bounds on the infection time delay between two persons w.r.t. their respective quarantine dates, conditioned on them not being part of the same household. Like Flaxman et al[7], the serial interval distribution, $s^*$, was chosen to be gamma distributed with the parameters

$$s^* \sim \Gamma(\alpha = 2.6, \beta = 0.4)$$

discretized by taking the density at each $t \in [1, \cdots, 100]$ and dividing by the sum over all $t$.

For each person, the infection time delay distribution was constrained based on the available data in the following way. Conditioned on the sampled infection time of person $i$, $t_{inf,i}$, we sample $\alpha_i$, the person that infected $i$. We define an upper bound on the time of infection from $\alpha_i$ to $i$ to be

$$t_{u,\alpha_i} = \begin{cases} \infty, & \text{if } \alpha_i \text{ was not quarantined at diagnosis,} \\ t_{r,i}, & \text{if } \alpha_i \text{ and } i \text{ share a household} \\ min\{t_{d,\alpha_i}, t_{q,\alpha_i}, t_{r,i}\}, & \text{otherwise.} \end{cases}$$

The likelihood $\mathcal{L}_{TD}$ of the delay $t = t_{inf,i}$- $t_{inf,\alpha_i}$ is as follows:

$$\mathcal{L}_{TI}(t) = \begin{cases} 0, & \text{if } t_{inf,i} > t_{u,\alpha_i} \\ \dfrac{s(t)}{\sum_{k=1}^{t_{u,\alpha_i}} s(k)}, & \text{otherwise.} \end{cases}$$

Thus, we allow for the possibility that a quarantined person to infect other members of their household, but no one outside of their household during quarantine.

##### Haplotype imputation

In the cases where an infected person had started producing antibodies at the time of sampling, the sequenced haplotype may have been incomplete or even missing altogether. If the sequence coverage was less than 95%, we chose a random person with whom they had reported contact and had them inherit their viral haplotype in the model.

##### Genetic likelihood

To estimate the likelihood of the viral haplotype of person $i$ conditioned on $\alpha_i$having infected them, we compare their set of mutations, $M_i$ and $M_{\alpha_i}$. Let $d_{m,i}$ be the sequencing depth over mutation $m$ in person $i$, measured as the number of reads overlapping a 1 base pair window around the start ($s_m$) and end site ($e_m$) of the mutation ($s_m = e_m$ if $m$ is a SNP or insertion). Further define $D_{i,\alpha_i} = \{m \in M_{\alpha_i} \setminus M_i : d_{m,i} < 5\}$ as the set of mutations in $\alpha_i$ with insufficient coverage in $i$. $D_{\alpha_i,i}$ is defined symmetrically to $D_{i,\alpha_i}$. We define the accumulation of mutations $r_{\alpha_i,i}$ from person $\alpha_i$ to $i$ as follows:

$$r_{\alpha_i,i} = \begin{cases} \left|(M_i \cup D_{i,\alpha_i}) \setminus M_{\alpha_i}\right|, & \text{if } M_{\alpha_i} \subseteq M_i \cup D_{i,\alpha_i}, \\ -\left|\left(M_{\alpha_i} \cup D_{\alpha_i,i}\right) \setminus M_i\right|, & \text{if } M_i \subset M_{\alpha_i} \cup D_{\alpha_i,i}, \\ -\infty, & \text{otherwise.} \end{cases}$$

Let $X$ be the random variable representing the accumulation of mutations between two persons separated by $\kappa$ generations of infections. We model $X$ with a Poisson random variable

$$X \sim Poi(\lambda = \kappa\mu),$$

where $\mu$ is the mutation rate. We define the genetic likelihood of the viral genotype of person $i$, conditioned on $\alpha_i$ having infected them

$$\mathcal{L}_G(M_i | \alpha_i, M_{\alpha_i}, \kappa_i, \mu) = P(X = r_{\alpha_i,i}).$$

Note that for $r_{\alpha_i,i} < 0$, $\mathcal{L}_G(M_i | \alpha_i, M_{\alpha_i}, \kappa_i, \mu) = 0$, which disallows the occurrence of back mutations and incompatible haplotypes and ignores sequencing errors.

##### Other likelihoods

The default Outbreaker2 contact and reporting likelihoods were used as described in Campbell et al[5].

#### Prior distributions

##### Estimate of the mutation rate of SARS-CoV-2

We estimated the accumulation of mutations in SARS-CoV-2 per transmission using pairs in the contact tracing network. We found 572 pairs of infected persons linked by contact tracing with a sequence coverage greater than 99%, that had their sampling dates separated by 2 days or more, where the receiving person in the pair shared haplotype or derived haplotype with the spreading person. The average number of mutations in the receiving persons that are not present in the spreading persons provides an estimate of the mutation rate per transmission. We found 158 such mutations among the 572 pairs, translating into a mutation rate of 0.28 per transmission. To correct for errors in the transmission chain inference and for false positive mutations, we reversed the role of the spreading and receiving person (459 pairs) and found 19 mutations (0.04 per transmission). This resulted in a corrected mutation rate of 0.23 per transmission (95%-CI: 0.19-0.28).

##### Other hyperparameters

The default Outbreaker2 prior distributions were used for the remaining parameters as described by Campbell et al[5]. The proportion of cases reported $\pi$, the proportion of contacts reported $\epsilon$, and the probability of non-infectious contact between cases $\lambda$ were all fitted by Outbreaker2. Posterior distributions of these hyperparameters are available in Supplementary table 5. The mutation rate of the virus $\mu$ was statically estimated and fixed at 0.23 (See above). The probability of contact between transmission pairs $\eta$, and the probability of false-positive reporting of contact $\zeta$ were fixed to $\eta = 1, \zeta = 0$, as is the default in Outbreaker2.

#### The effective reproduction number of a transmission tree

Let $N$ be the number of persons in a given transmission tree. Each person except for the root is infected by exactly one other person, who is also in the tree, thus has an in-degree of 1. The number of edges in the tree is then $N - 1$, and since the average $R$ is the average out-degree of the transmission tree we have that $R = 1 - \frac{1}{N}$.

#### Completeness of symptom and contact data

Each diagnosed person was contacted multiple times by telephone over the course of their infection by a team designated by the authorities. They recorded the onset and development of symptoms until 14 days or longer had passed since the last positive PCR and 7 days or longer had passed without symptoms. Furthermore, they recorded all persons the diagnosed person had been in contact with in the 48 hours prior to onset of symptoms or diagnosis, if they were asymptomatic at diagnosis, and the duration and intimacy of contact. All such contacts were required to go into quarantine. If the place of quarantine was shared with an infected person, the length of quarantine was two weeks, but otherwise a weeklong quarantine was sufficient. At the end of quarantine they were tested for SARS-CoV-2 regardless of whether they had developed symptoms or not.

#### Defining households

Using place of residence along with reported place of quarantine from the contact tracing data allowed us to group persons into distinct households. Infected persons sharing a household were deemed to have been able to infect one another, regardless of quarantine status (See Generation time likelihood). Using these households, we completed its contact tracing graph such that every person in a household had contact with every other person in the household.

#### Vaccination simulation sensitivity

In order to assess the sensitivity of the simulations to the initial cases in the tree, we repeated the simulations described in Main methods on subtrees of size greater than 100 with a root whose direct ancestor was one of the first 50 persons to be infected. A total of 1916 such subtrees were present in the 404 transmission trees that our model yielded. For each simulation we took the mean proportion of the initial size of the subtree (Supplementary figures 2-4).

#### Non-pharmaceutical interventions from September to December 1st 2020

Starting on August 19th, 2020, all persons entering Iceland were required to be PCR tested upon arrival. Given a negative test result, they had to quarantine for five days and tested again before leaving quarantine. Those who tested positive in either tested were required to isolate.

2021/09/11
Mandated quarantine shortened to 7 days from 14, and a negative PCR test required to leave quarantine.

2021/09/18
Bars and pubs closed for business.

2021/09/28
People required to remain seated while in restaurants.

2021/10/05
Social gatherings capped at 20 people, with some exceptions. Gyms closed. Public swimming pools open at 50% capacity.

2021/10/07
People required to stay 2 meters apart. Services that require a high level of proximity such as salons, beauty salons, massage parlors and tattoo studios closed. Customers and employees required to wear face masks in shops. Public swimming pools closed. Indoor sports practices forbidden. An audience of no more than 20 people allowed in theaters, movie theaters, and concerts. Restaurants required to close at 9 PM.

2021/10/20
No audience allowed at sports events. Gyms allowed to host classes at limited capacity and with thorough decontamination between classes. All children’s sports practices and after-school activities forbidden.

2021/10/31
Social gatherings capped at 10 people with some exceptions. All group sports practices forbidden. Theaters, movie theaters, and concert venues closed.

2021/11/03

Everyone older than 10 years old required to wear a mask and respect the 2 meter rule. Customers in grocery stores and pharmacies capped at 50 people and 10 people in other stores.

2021/11/18
Services that require a high level of proximity such as salons, beauty salons, massage parlors and tattoo studios allowed, but mask use mandatory. Children’s sport practices and after-school activities allowed with some restrictions.

#### Sequence data

##### MiSeq sequencing

The process used to obtain the sequences via MiSeq are detailed in[2].

##### RNA extraction

Viral RNA samples were extracted from swabs (96 samples per 40 min run) using the Chemagic Viral RNA kit on the Chemagic360 instrument from Perkin Elmer, with 300/100 µL input/output volume(s), respectively. Each step in the workflow was monitored using an in-house LIMS (VirLab) with 2D barcoding (Greiner, 300 µL tubes) of all extracted positive samples.

##### Reverse transcription and PCR amplification

Reverse transcription (RT) and multiplex PCR was performed based on information provided by the Artic Network initiative (https://artic.network/) to generate cDNA. In short, extracted viral RNA was pre-incubated at 65 °C for 5 min in the presence of random hexamers (2.5 µM) and dNTP´s (500 µM). Sample cooling on ice was then followed by RT using SuperScript IV (ThermoFisher) in the presence of DTT (5 mM) and RNaseOUT inhibitor (Thermo Fisher) for 10 min at 42°C, followed by 10 min at 70 °C. Multiplex PCR of the resulting SARS-CoV-2 cDNA was performed using a tiling scheme of primers, designed to generate overlapping amplicons of approximately 400 bp (Supplementary table 6). Two PCR reactions were performed for each sample using primer pools A and B, respectively (Table S1). PCR amplification was done using the Q5® Hot Start High-Fidelity polymerase (New England Biolabs) with primers at 1 µM concentration. The reactions were performed in an MJR thermal cycler with a heated lid at 105 °C, using 35 cycles of denaturation (15 sec at 98 °C) and annealing/extension (5 min at 65 °C). The resulting PCR amplicon pools A and B were combined, purified using 1X Ampure XP magnetic beads (Beckman Coulter) and quantified using the Qubit dsDNA high-sensitivity assay kit (Thermo Fisher).

#### Sample preparation for ONT Sequencing

Pooled amplicons (50-400 ng) were prepared for ONT sequencing using native barcoding with up to 24 barcodes. The first step in the procedure was end-repair using NEBNext Ultra II End Prep Reaction Buffer and Enzyme mix, respectively (NEB/E7546L). Samples (50 µL) were incubated in an MJR thermocycler at 20 °C for 10 min, followed by 10 min at 65 °C and cooling at 4 °C. Following AMPure XP clean-up (1X), the native barcodes (NBD104/NBD114) were ligated by mixing 20 µL purified sample, 2.5 µL barcode and 25 uL blunt/T4 ligase mastermix (NEB/M0367L). The mixture was incubated at room temperature on a rotator mixer for 10 min. Samples were purified by AMPure XP and quantified using the Qubit dsDNA high-sensitivity assay kit. Samples were pooled appropriately based on yield and/or Ct values from previous RT-qPCR measurements, with up to 24 samples/pool. Pooled DNA samples (up to 90 uL) were next incubated on a rotator mixer with 5 uL adapter mix II (AMII/ONT), 20 uL NEBNext Quick ligation reaction buffer and 10 uL Quick T4 DNA ligase (NEB/E6056L) for 10 min at room temperature followed by AMPure clean-up and bead washing with 2x120 µL of short fragment buffer (SFB/ONT). Final pooled sample libraries were resuspended in 35 uL of EB buffer and used immediately for sequencing.

#### ONT Sequencing

Pooled libraries were sequenced on either GridION X5 or PromethION sequencers, using the MinION (R9.4.1/FLO-MIN106D) and PromethION (R9.4.1/FLO-PRO002) flowcells, respectively. Following priming of the flowcells, the libraries were mixed with sequencing buffer (SQB) and loading beads (LB) and loaded onto the flowcells in the appropriate volume depending on flowcell type (75 µL or 150 µL). Sequencing runs were performed using on-board, high-accuracy basecalling (Guppy) with generation of FASTQ files for each barcode. Data acquisition varied from 1-4 hrs depending on the number of samples/pool and run performance. At least 100K reads were acquired per sample (barcode).

### Supplementary results

#### Undiagnosed cases

An average of 303 persons (12%) in the model had a number of transmissions between them and their ancestor greater than one, indicating the presence of undiagnosed cases (see also Supplementary table 5). An average of 255 had one hidden ancestor, 40 had two hidden ancestors, 7 had three hidden ancestors, and 2 had four hidden ancestors. Some of these 303 undiagnosed cases might refer to the same undiagnosed person, but this number does not include undiagnosed persons that did not infect anyone else, i.e. leaves in the transmission tree. If an undiagnosed person is a leaf, we underestimate the numerator of their ancestor's group's $\hat{R}$ by one and the denominator of their (latent) group $\hat{R}$ by one. If an undiagnosed person is not a leaf, we underestimate both the numerator and the denominator of the $\hat{R}$ of their (latent) group by one. The mean proportion of persons with at least one hidden ancestor in the growth phase was 0.09 (0.06-0.12) and the mean in the decline phase was 0.16 (0.12-0.22). The vaccination model does not take into account the undiagnosed cases. This is due to the fact that some of the undiagnosed cases may refer to the same person and furthermore, their age is unknown.

#### Time distributions

We calculated the generation time, the time from one inferred infection time to the next, for each transmission pair in each transmission tree. By averaging these distributions over the trees, we obtained a distribution that was different from the distribution in Flaxman et al., which was assumed a priori in the model (Supplementary methods), with more probability mass at shorter generation times (Supplementary figure 6). It is possible that the generation time under the stringent non-pharmaceutical interventions that were in place during the third wave was shorter than the generation time in an outbreak with less severe interventions, which would explain this discrepancy. For example, a high proportion of cases in the third wave were diagnosed while in quarantine, which means that if they infected others, it most likely occurred soon after exposure, before they were quarantined.
Furthermore, we calculated the incubation time, the time from infection to onset of symptoms and found that it is similar to the distribution in Flaxman et al. (Supplementary figure 6, Supplementary methods).

### Supplementary tables

| Group 1 | Group 2 | $\hat{R}$ 1 | $\hat{R}$ 2 | Effect | $p$ |
|---|---|---|---|---|---|
| Overall, outside quarantine | Overall, in quarantine | 1.31 (1.21-1.43) | 0.69 (0.66-0.73) | 0.69 (0.66-0.73) | $2.8x10^{-32}$ |
| Overall, outside quarantine | Overall, long quarantine | 1.31 (1.21-1.43) | 0.54 (0.50-0.58) | 0.54 (0.5-0.58) | $2.5x10^{-50}$ |
| Overall, short quarantine | Overall, long quarantine | 0.89 (0.83-0.96) | 0.54 (0.50-0.58) | 0.54 (0.5-0.58) | $4.0x10^{-19}$ |
| Overall, 16+ y.o. | Overall, 0-15 y.o. | 1.06 (0.98-1.12) | 0.66 (0.59-0.73) | 0.66 (0.59-0.73) | $1.8x10^{-12}$ |
| Overall, 16-66 y.o. | Overall, 0-15 y.o. | 1.08 (1.01-1.16) | 0.66 (0.59-0.74) | 0.66 (0.59-0.74) | $4.6x10^{-13}$ |
| Overall, 16-66 y.o. | Overall, Outside working age | 1.08 (1.01-1.16) | 0.74 (0.66-0.84) | 0.74 (0.66-0.84) | $1.6x10^{-8}$ |
| Overall, 16-66 y.o. | Overall, 67+ y.o. | 1.08 (1.01-1.16) | 0.85 (0.7-1.05) | 0.85 (0.7-1.05) | $3.0x10^{-2}$ |
| Growth phase, outside quarantine | Growth phase, in quarantine | 1.45 (1.32-1.62) | 0.84 (0.78-0.91) | 71.8% (51.8%-96.7%) | $2.7x10^{-16}$ |
| Growth phase, outside quarantine | Growth phase, long quarantine | 1.45 (1.32-1.6) | 0.66 (0.58-0.74) | 120.2% (90.6%-156.3%) | $1.2x10^{-24}$ |
| Growth phase, short quarantine | Growth phase, long quarantine | 1.02 (0.93-1.13) | 0.66 (0.58-0.74) | 55.1% (32.4%-79.1%) | $1.5x10^{-8}$ |
| Growth phase, 16+ y.o. | Growth phase, children 0-15 y.o. | 1.22 (1.13-1.32) | 0.80 (0.69-0.93) | 51.7% (28.6%-78%) | $5.0x10^{-7}$ |
| Growth phase, 16-66 y.o. | Growth phase, outside working age | 1.25 (1.15-1.37) | 0.83 (0.74-0.92) | 50.6% (32.5%-74.3%) | $1.4x10^{-8}$ |
| Growth phase, 16-66 y.o. | Growth phase, 67+ y.o. | 1.25 (1.15-1.36) | 0.87 (0.74-1.02) | 42.9% (21%-74.7%) | $1.0x10^{-4}$ |
| Decline phase, outside quarantine | Decline phase, in quarantine | 1.08 (0.93-1.25) | 0.53 (0.49-0.57) | 103.8% (73.9%-142.3%) | $4.4x10^{-16}$ |
| Decline phase, outside quarantine | Decline phase, long quarantine | 1.08 (0.92-1.27) | 0.43 (0.39-0.48) | 149.8% (104.1%-200.7%) | $3.7x10^{-20}$ |
| Decline phase, short quarantine | Decline phase, long quarantine | 0.70 (0.62-0.78) | 0.43 (0.39-0.48) | 61.4% (38.7%-87.7%) | $1.4x10^{-9}$ |
| Decline phase, 16+ y.o. | Decline phase, 0-15 y.o. | 0.82 (0.74-0.91) | 0.53 (0.45-0.62) | 55.5% (27.6%-90.7%) | $1.5x10^{-5}$ |
| Decline phase, 16-66 y.o. | Decline phase, outside working age | 0.82 (0.73-0.92) | 0.66 (0.54-0.81) | 23.8% (-3.4%-56.3%) | $8.0x10^{-2}$ |
| Decline phase, 16-66 y.o. | Decline phase, 67+ y.o. | 0.82 (0.74-0.92) | 0.83 (0.59-1.14) | -1.6% (-30.2%-40.6%) | 0.9 |
| Growth phase, everyone | Decline phase, everyone | 1.17 (1.09-1.27) | 0.77 (0.7-0.85) | 51.7% (33.1%-71.3%) | $1.9x10^{-10}$ |
| Growth phase, outside quarantine | Decline phase, outside quarantine | 1.45 (1.32-1.62) | 1.08 (0.93-1.25) | 34.0% (12.2%-60.9%) | $1.7x10^{-3}$ |
| Growth phase, in quarantine | Decline phase, in quarantine | 0.84 (0.78-0.91) | 0.53 (0.49-0.57) | 59.0% (42.4%-78.4%) | $1.0x10^{-15}$ |
| Growth phase, short quarantine | Decline phase, short quarantine | 1.02 (0.93-1.13) | 0.70 (0.62-0.78) | 46.2% (26.2%-69.6%) | $5.4x10^{-7}$ |
| Growth phase, long quarantine | Decline phase, long quarantine | 0.66 (0.58-0.74) | 0.43 (0.39-0.48) | 52.1% (29.9%-79%) | $1.8x10^{-7}$ |
| Growth phase, 16+ y.o. | Decline phase, 16+ y.o. | 1.22 (1.13-1.32) | 0.82 (0.74-0.91) | 48.5% (30.9%-69.5%) | $4.3x10^{-9}$ |
| Growth phase, 0-15 y.o. | Decline phase, 0-15 y.o. | 0.80 (0.69-0.93) | 0.53 (0.45-0.62) | 52.2% (22.9%-90.8%) | $1.9x10^{-4}$ |
| Growth phase, 16-66 y.o. | Decline phase, 16-66 y.o. | 1.25 (1.15-1.37) | 0.82 (0.73-0.92) | 52.5% (33.2%-75.5%) | $6.3x10^{-9}$ |
| Growth phase, outside working age | Decline phase, outside working age | 0.83 (0.74-0.92) | 0.66 (0.54-0.81) | 25.3% (0.2%-56.7%) | $6.0x10^{-2}$ |
| Growth phase, 67+ y.o. | Decline phase, 67+ y.o. | 0.87 (0.74-1.02) | 0.83 (0.59-1.14) | 5.0% (-27.1%-53.7%) | 0.8 |

***Supplementary table 1.*** Effect sizes and p-values for comparisons between various groups in the growth phase, decline phase, and overall.

| Locus | Ref | Alt |
|---|---|---|
| 241 | C | T |
| 445 | T | C |
| 3037 | C | T |
| 6023 | T | C |
| 6286 | C | T |
| 8017 | A | G |
| 13064 | C | T |
| 14408 | C | T |
| 18483 | T | C |
| 19999 | G | T |
| 20229 | C | T |
| 21255 | G | C |
| 22227 | C | T |
| 23403 | A | G |
| 25563 | G | T |
| 26801 | C | G |
| 28932 | C | T |
| 29645 | G | T |

***Supplementary table 2.*** The mutations that make up the blue clade, representing the vast majority of diagnosed cases in the third wave.

| Age | One dose | Two doses | Total | % |
|---|---|---|---|---|
| 16-29 | 533 | 369 | 902 | 1.2% |
| 30-39 | 591 | 443 | 1034 | 1.9% |
| 40-49 | 614 | 398 | 1012 | 2.1% |
| 50-59 | 568 | 358 | 926 | 2.1% |
| 60-69 | 608 | 480 | 1088 | 2.8% |
| 70-79 | 849 | 562 | 1411 | 5.8% |
| 80-89 | 1567 | 1253 | 2820 | 27.3% |
| 90+ | 599 | 963 | 1562 | 63.0% |

***Supplementary table 3***. Age composition of persons vaccinated by the end of the third wave, on January 28, 2021.

| Age | One dose | Two doses | Total | % |
|---|---|---|---|---|
| 16-29 | 5494 | 1961 | 7455 | 10.1% |
| 30-39 | 4969 | 2129 | 7098 | 13.0% |
| 40-49 | 6798 | 2241 | 9039 | 18.8% |
| 50-59 | 10955 | 2301 | 13256 | 30.4% |
| 60-69 | 21305 | 4464 | 25769 | 67.3% |
| 70-79 | 13290 | 9641 | 22931 | 95.0% |
| 80-89 | 150 | 9907 | 10057 | 97.5% |
| 90+ | 7 | 2412 | 2419 | 97.6% |

***Supplementary table 4***. Age composition of persons vaccinated by April 28, 2021.

| Parameter | Mean | 95%-PI |
|---|---|---|
| $\epsilon$ | 0.74 | (0.72 - 0.77) |
| $\lambda$ | $5.5 \times 10^{-4}$ | ($5.3 \times 10^{-4}$ – $5.9 \times 10^{-4}$) |
| $\pi$ | 0.87 | (0.83 - 0.91) |

***Supplementary table 5***. Posterior summary of the proportion of cases sampled $\pi$, the proportion of cases reported $\epsilon$, and the probability of non-infectious contact between cases $\lambda$. PI refers to the posterior interval.

| name | pool | seq | length | %gc | tm |
|---|---|---|---|---|---|
| nCoV-2019_1_LEFT | nCoV-2019_1 | ACCAACCAACTTTCGATCTCTTGT | 24 | 41.67 | 60.69 |
| nCoV-2019_1_RIGHT | nCoV-2019_1 | CATCTTTAAGATGTTGACGTGCCTC | 25 | 44 | 60.45 |
| nCoV-2019_2_LEFT | nCoV-2019_2 | CTGTTTTACAGGTTCGCGACGT | 22 | 50 | 61.67 |
| nCoV-2019_2_RIGHT | nCoV-2019_2 | TAAGGATCAGTGCCAAGCTCGT | 22 | 50 | 61.74 |
| nCoV-2019_3_LEFT | nCoV-2019_1 | CGGTAATAAAGGAGCTGGTGGC | 22 | 54.55 | 61.32 |
| nCoV-2019_3_RIGHT | nCoV-2019_1 | AAGGTGTCTGCAATTCATAGCTCT | 24 | 41.67 | 60.32 |
| nCoV-2019_4_LEFT | nCoV-2019_2 | GGTGTATACTGCTGCCGTGAAC | 22 | 54.55 | 61.56 |
| nCoV-2019_4_RIGHT | nCoV-2019_2 | CACAAGTAGTGGCACCTTCTTTAGT | 25 | 44 | 60.97 |
| nCoV-2019_5_LEFT | nCoV-2019_1 | TGGTGAAACTTCATGGCAGACG | 22 | 50 | 61.39 |
| nCoV-2019_5_RIGHT | nCoV-2019_1 | ATTGATGTTGACTTTCTCTTTTTGGAGT | 28 | 32.14 | 60.17 |
| nCoV-2019_6_LEFT | nCoV-2019_2 | GGTGTTGTTGGAGAAGGTTCCG | 22 | 54.55 | 61.64 |
| nCoV-2019_6_RIGHT | nCoV-2019_2 | TAGCGGCCTTCTGTAAAACACG | 22 | 50 | 61.18 |
| nCoV-2019_7_LEFT | nCoV-2019_1 | ATCAGAGGCTGCTCGTGTTGTA | 22 | 50 | 61.73 |
| nCoV-2019_7_RIGHT | nCoV-2019_1 | TGCACAGGTGACAATTTGTCCA | 22 | 45.45 | 60.95 |
| nCoV-2019_8_LEFT | nCoV-2019_2 | AGAGTTTCTTAGAGACGGTTGGGA | 24 | 45.83 | 61 |
| nCoV-2019_8_RIGHT | nCoV-2019_2 | GCTTCAACAGCTTCACTAGTAGGT | 24 | 45.83 | 60.56 |
| nCoV-2019_9_LEFT | nCoV-2019_1 | TCCCACAGAAGTGTTAACAGAGGA | 24 | 45.83 | 61.18 |
| nCoV-2019_9_RIGHT | nCoV-2019_1 | ATGACAGCATCTGCCACAACAC | 22 | 50 | 61.71 |
| nCoV-2019_10_LEFT | nCoV-2019_2 | TGAGAAGTGCTCTGCCTATACAGT | 24 | 45.83 | 61.12 |
| nCoV-2019_10_RIGHT | nCoV-2019_2 | TCATCTAACCAATCTTCTTCTTGCTCT | 27 | 37.04 | 60.31 |
| nCoV-2019_11_LEFT | nCoV-2019_1 | GGAATTTGGTGCCACTTCTGCT | 22 | 50 | 61.66 |
| nCoV-2019_11_RIGHT | nCoV-2019_1 | TCATCAGATTCAACTTGCATGGCA | 24 | 41.67 | 61.35 |
| nCoV-2019_12_LEFT | nCoV-2019_2 | AAACATGGAGGAGGTGTTGCAG | 22 | 50 | 61.08 |
| nCoV-2019_12_RIGHT | nCoV-2019_2 | TTCACTCTTCATTTCCAAAAAGCTTGA | 27 | 33.33 | 60.36 |
| nCoV-2019_13_LEFT | nCoV-2019_1 | TCGCACAAATGTCTACTTAGCTGT | 24 | 41.67 | 60.56 |
| nCoV-2019_13_RIGHT | nCoV-2019_1 | ACCACAGCAGTTAAAACACCCT | 22 | 45.45 | 60.36 |
| nCoV-2019_14_LEFT | nCoV-2019_2 | CATCCAGATTCTGCCACTCTTGT | 23 | 47.83 | 60.62 |
| nCoV-2019_14_RIGHT | nCoV-2019_2 | AGTTTCCACACAGACAGGCATT | 22 | 45.45 | 60.42 |
| nCoV-2019_15_LEFT | nCoV-2019_1 | ACAGTGCTTAAAAAGTGTAAAAGTGCC | 27 | 37.04 | 61.32 |
| nCoV-2019_15_RIGHT | nCoV-2019_1 | AACAGAAACTGTAGCTGGCACT | 22 | 45.45 | 60.16 |
| nCoV-2019_16_LEFT | nCoV-2019_2 | AATTTGGAAGAAGCTGCTCGGT | 22 | 45.45 | 60.82 |
| nCoV-2019_16_RIGHT | nCoV-2019_2 | CACAACTTGCGTGTGGAGGTTA | 22 | 50 | 61.32 |
| nCoV-2019_17_LEFT | nCoV-2019_1 | CTTCTTTCTTTGAGAGAAGTGAGGACT | 27 | 40.74 | 60.69 |
| nCoV-2019_17_RIGHT | nCoV-2019_1 | TTTGTTGGAGTGTTAACAATGCAGT | 25 | 36 | 60.11 |
| nCoV-2019_18_LEFT | nCoV-2019_2 | TGGAAATACCCACAAGTTAATGGTTTAAC | 29 | 34.48 | 60.69 |
| nCoV-2019_18_RIGHT | nCoV-2019_2 | AGCTTGTTTACCACACGTACAAGG | 24 | 45.83 | 61.51 |
| nCoV-2019_19_LEFT | nCoV-2019_1 | GCTGTTATGTACATGGGCACACT | 23 | 47.83 | 61.18 |
| nCoV-2019_19_RIGHT | nCoV-2019_1 | TGTCCAACTTAGGGTCAATTTCTGT | 25 | 40 | 60.4 |
| nCoV-2019_20_LEFT | nCoV-2019_2 | ACAAAGAAAACAGTTACACAACAACCA | 27 | 33.33 | 60.68 |
| nCoV-2019_20_RIGHT | nCoV-2019_2 | ACGTGGCTTTATTAGTTGCATTGTT | 25 | 36 | 60.28 |
| nCoV-2019_21_LEFT | nCoV-2019_1 | TGGCTATTGATTATAAACACTACACACCC | 29 | 37.93 | 61.49 |
| nCoV-2019_21_RIGHT | nCoV-2019_1 | TAGATCTGTGTGGCCAACCTCT | 22 | 50 | 60.83 |
| nCoV-2019_22_LEFT | nCoV-2019_2 | ACTACCGAAGTTGTAGGAGACATTATACT | 29 | 37.93 | 61.25 |
| nCoV-2019_22_RIGHT | nCoV-2019_2 | ACAGTATTCTTTGCTATAGTAGTCGGC | 27 | 40.74 | 60.73 |
| nCoV-2019_23_LEFT | nCoV-2019_1 | ACAACTACTAACATAGTTACACGGTGT | 27 | 37.04 | 60.26 |
| nCoV-2019_23_RIGHT | nCoV-2019_1 | ACCAGTACAGTAGGTTGCAATAGTG | 25 | 44 | 60.57 |
| nCoV-2019_24_LEFT | nCoV-2019_2 | AGGCATGCCTTCTTACTGTACTG | 23 | 47.83 | 60.37 |
| nCoV-2019_24_RIGHT | nCoV-2019_2 | ACATTCTAACCATAGCTGAAATCGGG | 26 | 42.31 | 61.19 |
| nCoV-2019_25_LEFT | nCoV-2019_1 | GCAATTGTTTTTCAGCTATTTTGCAGT | 27 | 33.33 | 60.73 |
| nCoV-2019_25_RIGHT | nCoV-2019_1 | ACTGTAGTGACAAGTCTCTCGCA | 23 | 47.83 | 61.3 |
| nCoV-2019_26_LEFT | nCoV-2019_2 | TTGTGATACATTCTGTGCTGGTAGT | 25 | 40 | 60.28 |
| nCoV-2019_26_RIGHT | nCoV-2019_2 | TCCGCACTATCACCAACATCAG | 22 | 50 | 60.42 |
| nCoV-2019_27_LEFT | nCoV-2019_1 | ACTACAGTCAGCTTATGTGTCAACC | 25 | 44 | 60.8 |
| nCoV-2019_27_RIGHT | nCoV-2019_1 | AATACAAGCACCAAGGTCACGG | 22 | 50 | 61.13 |
| nCoV-2019_28_LEFT | nCoV-2019_2 | ACATAGAAGTTACTGGCGATAGTTGT | 26 | 38.46 | 60.13 |
| nCoV-2019_28_RIGHT | nCoV-2019_2 | TGTTTAGACATGACATGAACAGGTGT | 26 | 38.46 | 60.91 |
| nCoV-2019_29_LEFT | nCoV-2019_1 | ACTTGTGTTCCTTTTTGTTGCTGC | 24 | 41.67 | 61.39 |
| nCoV-2019_29_RIGHT | nCoV-2019_1 | AGTGTACTCTATAAGTTTTGATGGTGTGT | 29 | 34.48 | 60.69 |
| nCoV-2019_30_LEFT | nCoV-2019_2 | GCACAACTAATGGTGACTTTTTGCA | 25 | 40 | 61.19 |
| nCoV-2019_30_RIGHT | nCoV-2019_2 | ACCACTAGTAGATACACAAACACCAG | 26 | 42.31 | 60.3 |
| nCoV-2019_31_LEFT | nCoV-2019_1 | TTCTGAGTACTGTAGGCACGGC | 22 | 54.55 | 62.03 |
| nCoV-2019_31_RIGHT | nCoV-2019_1 | ACAGAATAAACACCAGGTAAGAATGAGT | 28 | 35.71 | 60.69 |
| nCoV-2019_32_LEFT | nCoV-2019_2 | TGGTGAATACAGTCATGTAGTTGCC | 25 | 44 | 61.09 |
| nCoV-2019_32_RIGHT | nCoV-2019_2 | AGCACATCACTACGCAACTTTAGA | 24 | 41.67 | 60.56 |
| nCoV-2019_33_LEFT | nCoV-2019_1 | ACTTTTGAAGAAGCTGCGCTGT | 22 | 45.45 | 61.58 |
| nCoV-2019_33_RIGHT | nCoV-2019_1 | TGGACAGTAAACTACGTCATCAAGC | 25 | 44 | 61.08 |
| nCoV-2019_34_LEFT | nCoV-2019_2 | TCCCATCTGGTAAAGTTGAGGGT | 23 | 47.83 | 61.02 |
| nCoV-2019_34_RIGHT | nCoV-2019_2 | AGTGAAATTGGGCCTCATAGCA | 22 | 45.45 | 60.03 |
| nCoV-2019_35_LEFT | nCoV-2019_1 | TGTTCGCATTCAACCAGGACAG | 22 | 50 | 61.39 |
| nCoV-2019_35_RIGHT | nCoV-2019_1 | ACTTCATAGCCACAAGGTTAAAGTCA | 26 | 38.46 | 60.69 |
| nCoV-2019_36_LEFT | nCoV-2019_2 | TTAGCTTGGTTGTACGCTGCTG | 22 | 50 | 61.44 |
| nCoV-2019_36_RIGHT | nCoV-2019_2 | GAACAAAGACCATTGAGTACTCTGGA | 26 | 42.31 | 60.74 |

| | | | | | |
|---|---|---|---|---|---|
| nCoV-2019_37_LEFT | nCoV-2019_1 | ACACACCACTGGTTGTTACTCAC | 23 | 47.83 | 60.93 |
| nCoV-2019_37_RIGHT | nCoV-2019_1 | GTCCACACTCTCCTAGCACCAT | 22 | 54.55 | 61.48 |
| nCoV-2019_38_LEFT | nCoV-2019_2 | ACTGTGTTATGTATGCATCAGCTGT | 25 | 40 | 60.86 |
| nCoV-2019_38_RIGHT | nCoV-2019_2 | CACCAAGAGTCAGTCTAAAGTAGCG | 25 | 48 | 61.13 |
| nCoV-2019_39_LEFT | nCoV-2019_1 | AGTATTGCCCTATTTTCTTCATAACTGGT | 29 | 34.48 | 61 |
| nCoV-2019_39_RIGHT | nCoV-2019_1 | TGTAACTGGACACATTGAGCCC | 22 | 50 | 60.55 |
| nCoV-2019_40_LEFT | nCoV-2019_2 | TGCACATCAGTAGTCTTACTCTCAGT | 26 | 42.31 | 61.25 |
| nCoV-2019_40_RIGHT | nCoV-2019_2 | CATGGCTGCATCACGGTCAAAT | 22 | 50 | 62.09 |
| nCoV-2019_41_LEFT | nCoV-2019_1 | GTTCCCTTCCATCATATGCAGCT | 23 | 47.83 | 60.75 |
| nCoV-2019_41_RIGHT | nCoV-2019_1 | TGGTATGACAACCATTAGTTTGGCT | 25 | 40 | 60.75 |
| nCoV-2019_42_LEFT | nCoV-2019_2 | TGCAAGAGATGGTTGTGTTCCC | 22 | 50 | 61.08 |
| nCoV-2019_42_RIGHT | nCoV-2019_2 | CCTACCTCCCTTTGTTGTGTTGT | 23 | 47.83 | 60.69 |
| nCoV-2019_43_LEFT | nCoV-2019_1 | TACGACAGATGTCTTGTGCTGC | 22 | 50 | 60.93 |
| nCoV-2019_43_RIGHT | nCoV-2019_1 | AGCAGCATCTACAGCAAAAGCA | 22 | 45.45 | 61.14 |
| nCoV-2019_44_LEFT | nCoV-2019_2 | TGCCACAGTACGTCTACAAGCT | 22 | 50 | 61.66 |
| nCoV-2019_44_RIGHT | nCoV-2019_2 | AACCTTTCCACATACCGCAGAC | 22 | 50 | 60.87 |
| nCoV-2019_45_LEFT | nCoV-2019_1 | TACCTACAACTTGTGCTAATGACCC | 25 | 44 | 60.57 |
| nCoV-2019_45_RIGHT | nCoV-2019_1 | AAATTGTTTCTTCATGTTGGTAGTTAGAGA | 30 | 30 | 60.01 |
| nCoV-2019_46_LEFT | nCoV-2019_2 | TGTCGCTTCCAAGAAAAGGACG | 22 | 50 | 61.38 |
| nCoV-2019_46_RIGHT | nCoV-2019_2 | CACGTTCACCTAAGTTGGCGTA | 22 | 50 | 60.86 |
| nCoV-2019_47_LEFT | nCoV-2019_1 | AGGACTGGTATGATTTTGTAGAAAACCC | 28 | 39.29 | 61.42 |
| nCoV-2019_47_RIGHT | nCoV-2019_1 | AATAACGGTCAAAGAGTTTTAACCTCTC | 28 | 35.71 | 60.06 |
| nCoV-2019_48_LEFT | nCoV-2019_2 | TGTTGACACTGACTTAACAAAGCCT | 25 | 40 | 61.09 |
| nCoV-2019_48_RIGHT | nCoV-2019_2 | TAGATTACCAGAAGCAGCGTGC | 22 | 50 | 60.74 |
| nCoV-2019_49_LEFT | nCoV-2019_1 | AGGAATTACTTGTGTATGCTGCTGA | 25 | 40 | 60.57 |
| nCoV-2019_49_RIGHT | nCoV-2019_1 | TGACGATGACTTGGTTAGCATTAATACA | 28 | 35.71 | 61.05 |
| nCoV-2019_50_LEFT | nCoV-2019_2 | GTTGATAAGTACTTTGATTGTTACGATGGT | 30 | 33.33 | 60.59 |
| nCoV-2019_50_RIGHT | nCoV-2019_2 | TAACATGTTGTGCCAACCACCA | 22 | 45.45 | 60.95 |
| nCoV-2019_51_LEFT | nCoV-2019_1 | TCAATAGCCGCCACTAGAGGAG | 22 | 54.55 | 61.34 |
| nCoV-2019_51_RIGHT | nCoV-2019_1 | AGTGCATTAACATTGGCCGTGA | 22 | 45.45 | 61.14 |
| nCoV-2019_52_LEFT | nCoV-2019_2 | CATCAGGAGATGCCACAACTGC | 22 | 54.55 | 61.83 |
| nCoV-2019_52_RIGHT | nCoV-2019_2 | GTTGAGAGCAAAATTCATGAGGTCC | 25 | 44 | 60.62 |
| nCoV-2019_53_LEFT | nCoV-2019_1 | AGCAAAATGTTGGACTGAGACTGA | 24 | 41.67 | 60.69 |
| nCoV-2019_53_RIGHT | nCoV-2019_1 | AGCCTCATAAAACTCAGGTTCCC | 23 | 47.83 | 60.31 |
| nCoV-2019_54_LEFT | nCoV-2019_2 | TGAGTTAACAGGACACATGTTAGACA | 26 | 38.46 | 60.18 |
| nCoV-2019_54_RIGHT | nCoV-2019_2 | AACCAAAAACTTGTCCATTAGCACA | 25 | 36 | 60.11 |
| nCoV-2019_55_LEFT | nCoV-2019_1 | ACTCAACTTTACTTAGGAGGTATGAGCT | 28 | 39.29 | 61.43 |
| nCoV-2019_55_RIGHT | nCoV-2019_1 | GGTGTACTCTCCTATTTGTACTTTACTGT | 29 | 37.93 | 60.54 |
| nCoV-2019_56_LEFT | nCoV-2019_2 | ACCTAGACCACCACTTAACCGA | 22 | 50 | 60.49 |
| nCoV-2019_56_RIGHT | nCoV-2019_2 | ACACTATGCGAGCAGAAGGGTA | 22 | 50 | 61.21 |
| nCoV-2019_57_LEFT | nCoV-2019_1 | ATTCTACACTCCAGGGACCACC | 22 | 54.55 | 61.16 |
| nCoV-2019_57_RIGHT | nCoV-2019_1 | GTAATTGAGCAGGGTCGCCAAT | 22 | 50 | 61.26 |
| nCoV-2019_58_LEFT | nCoV-2019_2 | TGATTTGAGTGTTGTCAATGCCAGA | 25 | 40 | 61.44 |
| nCoV-2019_58_RIGHT | nCoV-2019_2 | CTTTTCTCCAAGCAGGGTTACGT | 23 | 47.83 | 61.06 |
| nCoV-2019_59_LEFT | nCoV-2019_1 | TCACGCATGATGTTTCATCTGCA | 23 | 43.48 | 61.42 |
| nCoV-2019_59_RIGHT | nCoV-2019_1 | AAGAGTCCTGTTACATTTTCAGCTTG | 26 | 38.46 | 60.02 |
| nCoV-2019_60_LEFT | nCoV-2019_2 | TGATAGAGACCTTTATGACAAGTTGCA | 27 | 37.04 | 60.53 |
| nCoV-2019_60_RIGHT | nCoV-2019_2 | GGTACCAACAGCTTCTCTAGTAGC | 24 | 50 | 60.44 |
| nCoV-2019_61_LEFT | nCoV-2019_1 | TGTTTATCACCCGCGAAGAAGC | 22 | 50 | 61.5 |
| nCoV-2019_61_RIGHT | nCoV-2019_1 | ATCACATAGACAACAGGTGCGC | 22 | 50 | 61.25 |
| nCoV-2019_62_LEFT | nCoV-2019_2 | GGCACATGGCTTTGAGTTGACA | 22 | 50 | 61.91 |
| nCoV-2019_62_RIGHT | nCoV-2019_2 | GTTGAACCTTTCTACAAGCCGC | 22 | 50 | 60.35 |
| nCoV-2019_63_LEFT | nCoV-2019_1 | TGTTAAGCGTGTTGACTGGACT | 22 | 45.45 | 60.16 |
| nCoV-2019_63_RIGHT | nCoV-2019_1 | ACAAACTGCCACCATCACAACC | 22 | 50 | 61.85 |
| nCoV-2019_64_LEFT | nCoV-2019_2 | TCGATAGATATCCTGCTAATTCCATTGT | 28 | 35.71 | 60.11 |
| nCoV-2019_64_RIGHT | nCoV-2019_2 | AGTCTTGTAAAAGTGTTCCAGAGGT | 25 | 40 | 60.1 |
| nCoV-2019_65_LEFT | nCoV-2019_1 | GCTGGCTTTAGCTTGTGGGTTT | 22 | 50 | 61.92 |
| nCoV-2019_65_RIGHT | nCoV-2019_1 | TGTCAGTCATAGAACAAACACCAATAGT | 28 | 35.71 | 60.9 |
| nCoV-2019_66_LEFT | nCoV-2019_2 | GGGTGTGGACATTGCTGCTAAT | 22 | 50 | 61.21 |
| nCoV-2019_66_RIGHT | nCoV-2019_2 | TCAATTTCCATTTGACTCCTGGGT | 24 | 41.67 | 60.45 |
| nCoV-2019_67_LEFT | nCoV-2019_1 | GTTGTCCAACAATTACCTGAAACTTACT | 28 | 35.71 | 60.43 |
| nCoV-2019_67_RIGHT | nCoV-2019_1 | CAACCTTAGAAACTACAGATAAATCTTGGG | 30 | 36.67 | 60.4 |
| nCoV-2019_68_LEFT | nCoV-2019_2 | ACAGGTTCATCTAAGTGTGTGTGT | 24 | 41.67 | 60.14 |
| nCoV-2019_68_RIGHT | nCoV-2019_2 | CTCCTTTATCAGAACCAGCACCA | 23 | 47.83 | 60.31 |
| nCoV-2019_69_LEFT | nCoV-2019_1 | TGTCGCAAAATATACTCAACTGTGTCA | 27 | 37.04 | 61.43 |
| nCoV-2019_69_RIGHT | nCoV-2019_1 | TCTTTATAGCCACGGAACCTCCA | 23 | 47.83 | 61.14 |
| nCoV-2019_70_LEFT | nCoV-2019_2 | ACAAAAGAAAATGACTCTAAAGAGGGTTT | 29 | 31.03 | 60.13 |
| nCoV-2019_70_RIGHT | nCoV-2019_2 | TGACCTTCTTTTAAAGACATAACAGCAG | 28 | 35.71 | 60.27 |
| nCoV-2019_71_LEFT | nCoV-2019_1 | ACAAATCCAATTCAGTTGTCTTCCTATTC | 29 | 34.48 | 60.54 |
| nCoV-2019_71_RIGHT | nCoV-2019_1 | TGGAAAAGAAAGGTAAGAACAAGTCCT | 27 | 37.04 | 60.8 |
| nCoV-2019_72_LEFT | nCoV-2019_2 | ACACGTGGTGTTTATTACCCTGAC | 24 | 45.83 | 61.04 |
| nCoV-2019_72_RIGHT | nCoV-2019_2 | ACTCTGAACTCACTTTCCATCCAAC | 25 | 44 | 60.97 |
| nCoV-2019_73_LEFT | nCoV-2019_1 | CAATTTTGTAATGATCCATTTTTGGGTGT | 29 | 31.03 | 60.29 |
| nCoV-2019_73_RIGHT | nCoV-2019_1 | CACCAGCTGTCCAACCTGAAGA | 22 | 54.55 | 62.45 |
| nCoV-2019_74_LEFT | nCoV-2019_2 | ACATCACTAGGTTTCAAACTTTACTTGC | 28 | 35.71 | 60.68 |

| | | | | | |
|---|---|---|---|---|---|
| nCoV-2019_74_RIGHT | nCoV-2019_2 | GCAACACAGTTGCTGATTCTCTTC | 24 | 45.83 | 60.85 |
| nCoV-2019_75_LEFT | nCoV-2019_1 | AGAGTCCAACCAACAGAATCTATTGT | 26 | 38.46 | 60.24 |
| nCoV-2019_75_RIGHT | nCoV-2019_1 | ACCACCAACCTTAGAATCAAGATTGT | 26 | 38.46 | 60.69 |
| nCoV-2019_76_LEFT | nCoV-2019_2 | AGGGCAAACTGGAAAGATTGCT | 22 | 45.45 | 60.76 |
| nCoV-2019_76_RIGHT | nCoV-2019_2 | ACACCTGTGCCTGTTAAACCAT | 22 | 45.45 | 60.42 |
| nCoV-2019_77_LEFT | nCoV-2019_1 | CCAGCAACTGTTTGTGGACCTA | 22 | 50 | 60.75 |
| nCoV-2019_77_RIGHT | nCoV-2019_1 | CAGCCCCTATTAAACAGCCTGC | 22 | 54.55 | 61.59 |
| nCoV-2019_78_LEFT | nCoV-2019_2 | CAACTTACTCCTACTTGGCGTGT | 23 | 47.83 | 60.55 |
| nCoV-2019_78_RIGHT | nCoV-2019_2 | TGTGTACAAAAACTGCCATATTGCA | 25 | 36 | 60.22 |
| nCoV-2019_79_LEFT | nCoV-2019_1 | GTGGTGATTCAACTGAATGCAGC | 23 | 47.83 | 60.92 |
| nCoV-2019_79_RIGHT | nCoV-2019_1 | CATTTCATCTGTGAGCAAAGGTGG | 24 | 45.83 | 60.62 |
| nCoV-2019_80_LEFT | nCoV-2019_2 | TTGCCTTGGTGATATTGCTGCT | 22 | 45.45 | 60.89 |
| nCoV-2019_80_RIGHT | nCoV-2019_2 | TGGAGCTAAGTTGTTTAACAAGCG | 24 | 41.67 | 60.02 |
| nCoV-2019_81_LEFT | nCoV-2019_1 | GCACTTGGAAAACTTCAAGATGTGG | 25 | 44 | 61.24 |
| nCoV-2019_81_RIGHT | nCoV-2019_1 | GTGAAGTTCTTTTCTTGTGCAGGG | 24 | 45.83 | 60.73 |
| nCoV-2019_82_LEFT | nCoV-2019_2 | GGGCTATCATCTTATGTCCTTCCCT | 25 | 48 | 61.52 |
| nCoV-2019_82_RIGHT | nCoV-2019_2 | TGCCAGAGATGTCACCTAAATCAA | 24 | 41.67 | 60.02 |
| nCoV-2019_83_LEFT | nCoV-2019_1 | TCCTTTGCAACCTGAATTAGACTCA | 25 | 40 | 60.46 |
| nCoV-2019_83_RIGHT | nCoV-2019_1 | TTTGACTCCTTTGAGCACTGGC | 22 | 50 | 61.33 |
| nCoV-2019_84_LEFT | nCoV-2019_2 | TGCTGTAGTTGTCTCAAGGGCT | 22 | 50 | 61.61 |
| nCoV-2019_84_RIGHT | nCoV-2019_2 | AGGTGTGAGTAAACTGTTACAAACAAC | 27 | 37.04 | 60.36 |
| nCoV-2019_85_LEFT | nCoV-2019_1 | ACTAGCACTCTCCAAGGGTGTT | 22 | 50 | 61.03 |
| nCoV-2019_85_RIGHT | nCoV-2019_1 | ACACAGTCTTTTACTCCAGATTCCC | 25 | 44 | 60.51 |
| nCoV-2019_86_LEFT | nCoV-2019_2 | TCAGGTGATGGCACAACAAGTC | 22 | 50 | 61.07 |
| nCoV-2019_86_RIGHT | nCoV-2019_2 | ACGAAAGCAAGAAAAAGAAGTACGC | 25 | 40 | 61.01 |
| nCoV-2019_87_LEFT | nCoV-2019_1 | CGACTACTAGCGTGCCTTTGTA | 22 | 50 | 60.16 |
| nCoV-2019_87_RIGHT | nCoV-2019_1 | ACTAGGTTCCATTGTTCAAGGAGC | 24 | 45.83 | 60.81 |
| nCoV-2019_88_LEFT | nCoV-2019_2 | CCATGGCAGATTCCAACGGTAC | 22 | 54.55 | 61.58 |
| nCoV-2019_88_RIGHT | nCoV-2019_2 | TGGTCAGAATAGTGCCATGGAGT | 23 | 47.83 | 61.4 |
| nCoV-2019_89_LEFT | nCoV-2019_1 | GTACGCGTTCCATGTGGTCATT | 22 | 50 | 61.5 |
| nCoV-2019_89_RIGHT | nCoV-2019_1 | ACCTGAAAGTCAACGAGATGAAACA | 25 | 40 | 60.91 |
| nCoV-2019_90_LEFT | nCoV-2019_2 | ACACAGACCATTCCAGTAGCAGT | 23 | 47.83 | 61.58 |
| nCoV-2019_90_RIGHT | nCoV-2019_2 | TGAAATGGTGAATTGCCCTCGT | 22 | 45.45 | 60.82 |
| nCoV-2019_91_LEFT | nCoV-2019_1 | TCACTACCAAGAGTGTGTTAGAGGT | 25 | 44 | 60.93 |
| nCoV-2019_91_RIGHT | nCoV-2019_1 | TTCAAGTGAGAACCAAAAGATAATAAGCA | 29 | 31.03 | 60.03 |
| nCoV-2019_92_LEFT | nCoV-2019_2 | TTTGTGCTTTTTAGCCTTTCTGCT | 24 | 37.5 | 60.14 |
| nCoV-2019_92_RIGHT | nCoV-2019_2 | AGGTTCCTGGCAATTAATTGTAAAAGG | 27 | 37.04 | 60.53 |
| nCoV-2019_93_LEFT | nCoV-2019_1 | TGAGGCTGGTTCTAAATCACCCA | 23 | 47.83 | 61.59 |
| nCoV-2019_93_RIGHT | nCoV-2019_1 | AGGTCTTCCTTGCCATGTTGAG | 22 | 50 | 60.55 |
| nCoV-2019_94_LEFT | nCoV-2019_2 | GGCCCCAAGGTTTACCCAATAA | 22 | 50 | 60.56 |
| nCoV-2019_94_RIGHT | nCoV-2019_2 | TTTGGCAATGTTGTTCCTTGAGG | 23 | 43.48 | 60.18 |
| nCoV-2019_95_LEFT | nCoV-2019_1 | TGAGGGAGCCTTGAATACACCA | 22 | 50 | 61.1 |
| nCoV-2019_95_RIGHT | nCoV-2019_1 | CAGTACGTTTTTGCCGAGGCTT | 22 | 50 | 61.95 |
| nCoV-2019_96_LEFT | nCoV-2019_2 | GCCAACAACAACAAGGCCAAAC | 22 | 50 | 61.82 |
| nCoV-2019_96_RIGHT | nCoV-2019_2 | TAGGCTCTGTTGGTGGGAATGT | 22 | 50 | 61.36 |
| nCoV-2019_97_LEFT | nCoV-2019_1 | TGGATGACAAAGATCCAAATTTCAAAGA | 28 | 32.14 | 60.22 |
| nCoV-2019_97_RIGHT | nCoV-2019_1 | ACACACTGATTAAAGATTGCTATGTGAG | 28 | 35.71 | 60.17 |
| nCoV-2019_98_LEFT | nCoV-2019_2 | AACAATTGCAACAATCCATGAGCA | 24 | 37.5 | 60.5 |
| nCoV-2019_98_RIGHT | nCoV-2019_2 | TTCTCCTAAGAAGCTATTAAAATCACATGG | 30 | 33.33 | 60.01 |

***Supplementary table 6***. Amplicons used in ONT sequencing of SARS-CoV-2.

#### Supplementary figures

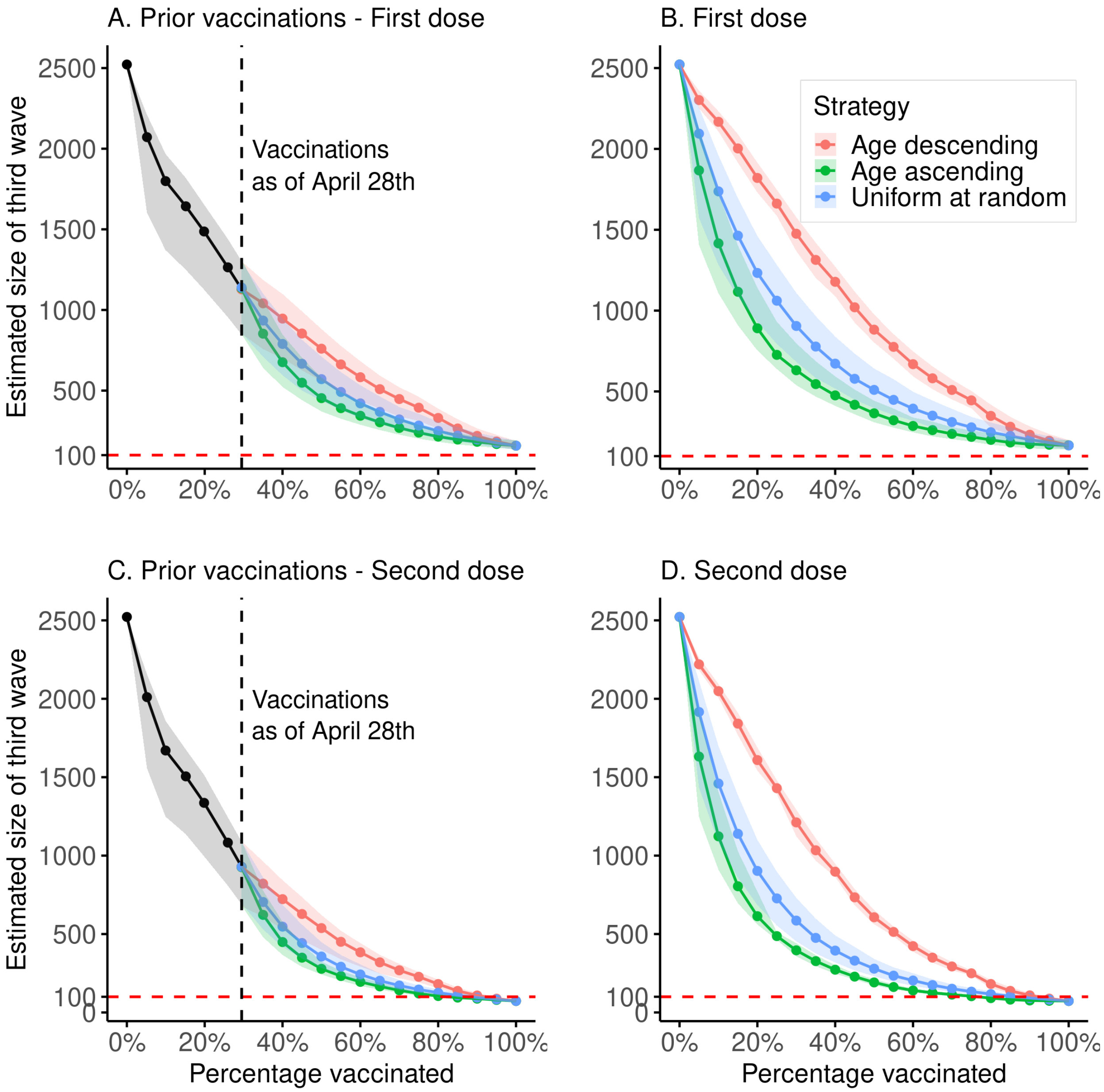


***Supplementary figure 1.*** Simulations of the estimated size of the third wave for a given population prevalence of vaccination, conditioned on the first 50 persons infected being unvaccinated. Solid lines show the mean outbreak size, shaded areas represent 2.5%-97.5% quantiles. **A**. Using the de facto vaccination scheme for at-risk groups and front-line workers, up to 29% of the adult population, and using three separate vaccination strategies from 29% to 100%: age-descending, age-ascending and uniformly at random. Modeled vaccinations beyond the 29% mark are assumed to have an efficacy of 60%. **B**. Simulations of the size of the third wave, assuming 60% vaccine efficacy, under the three different vaccination strategies, starting with no vaccinations and concluding with 100% of the adult population vaccinated. **C**. Same simulation as in A, but all vaccinations are assumed to have an efficacy of 90% (both first and second dose administered). **D**. Same simulation as in C, but assuming 90% vaccine efficacy.

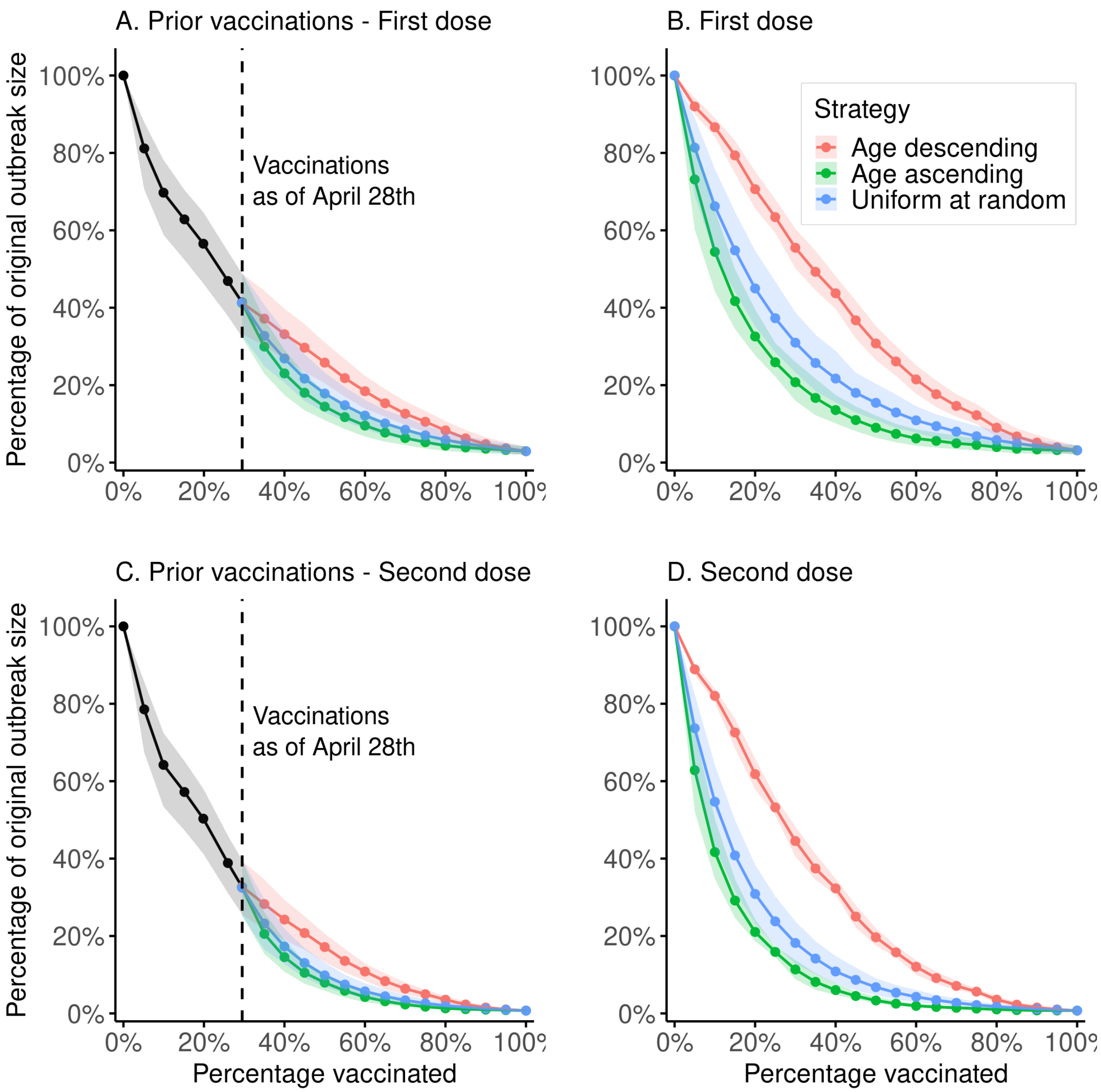


***Supplementary figure 2.*** Simulations of the estimated size of the third wave for a given population prevalence of vaccination. Each subtree is of size 100 and its root has a direct ancestor in the first 50 persons to be infected. Solid lines show the mean outbreak size, shaded areas represent 2.5%-97.5% quantiles. **A**. Using the de facto vaccination scheme for at-risk groups and front-line workers, up to 29% of the adult population, and using three separate vaccination strategies from 29% to 100%: age-descending, age-ascending and uniformly at random. Modeled vaccinations beyond the 29% mark are assumed to have an efficacy of 60%. **B**. Simulations of the size of the third wave, assuming 60% vaccine efficacy, under the three different vaccination strategies, starting with no vaccinations and concluding with 100% of the adult population vaccinated. **C**. Same simulation as in A, but all vaccinations are assumed to have an efficacy of 90% (both first and second dose administered). **D**. Same simulation as in C, but assuming 90% vaccine efficacy.

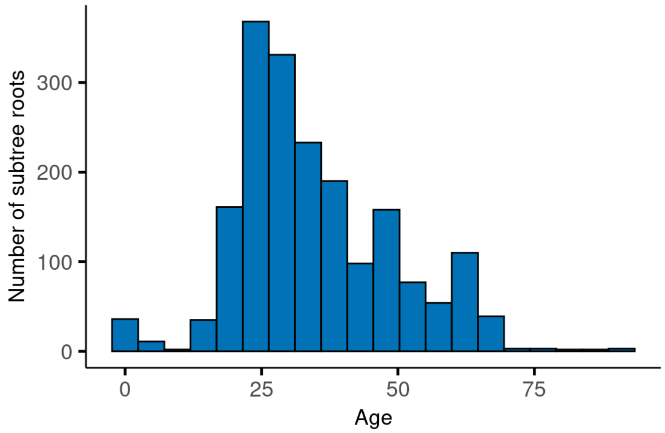


***Supplementary figure 3***. Age distribution of the roots in the subtrees in Supplementary figure 2.

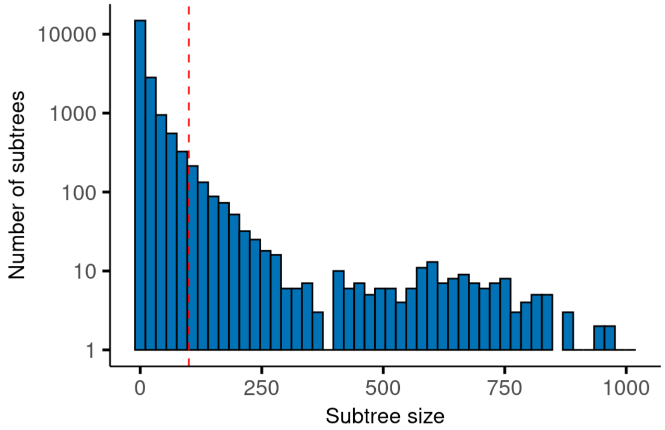


***Supplementary figure 4***. Log of the size distribution of the subtrees. The ones used in the simulations in Supplementary figure 2 are of size 100 or more (red, dotted line).

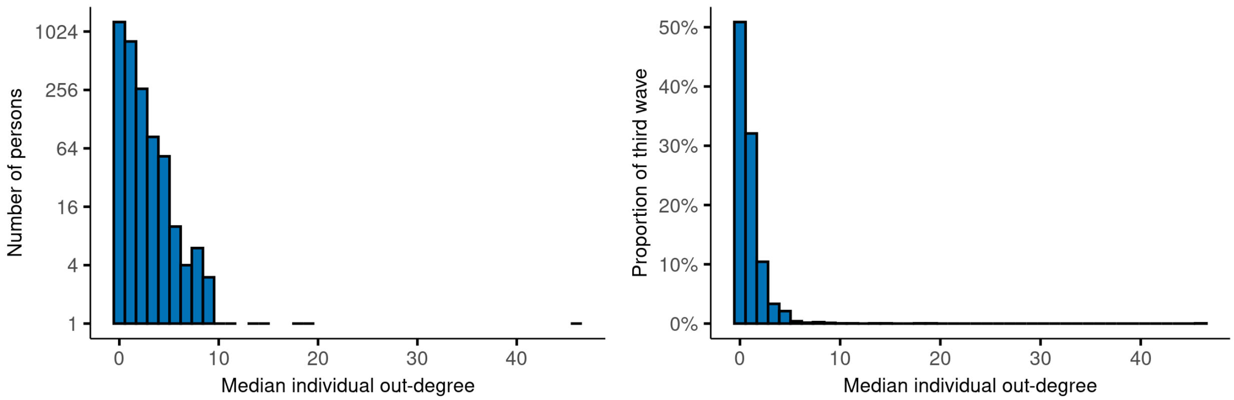


***Supplementary figure 5***. The distribution of the median out-degree of each person. The figure on the left shows the absolute number of persons in each bin on a log scale and the figure on the right shows the proportion of the third wave in each bin.

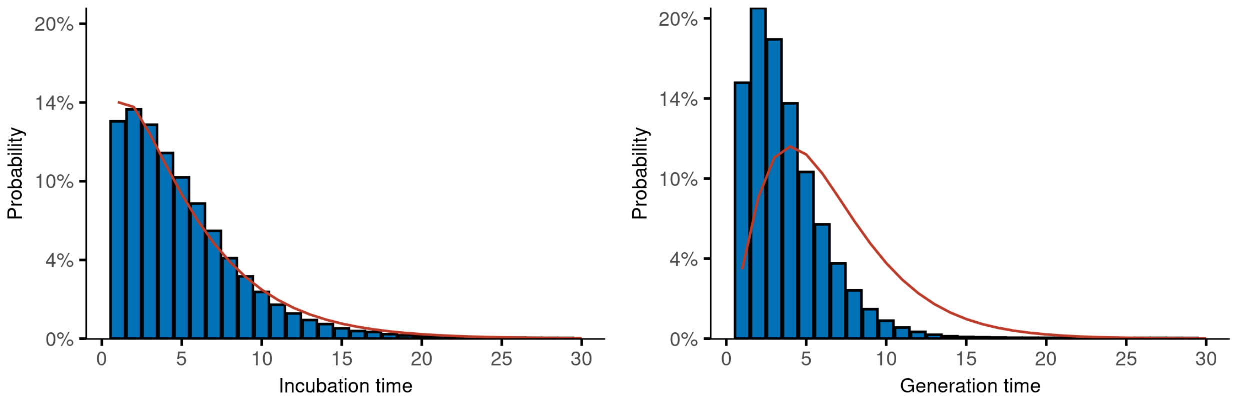


***Supplementary figure 6***. The incubation time distribution and generation time distribution averaged over all transmission trees. The red line shows the assumed distributions that were fed into the model[7]. The incubation time distribution refers to only those who were symptomatic.
